# Ethnic Inequalities in COVID-19 Vaccination Uptake Among Older Adults in Australia: A Nationwide Linked Data Study

**DOI:** 10.64898/2026.05.03.26351251

**Authors:** Peiyao Xu, Saman Khalatbari-Soltani, Meru Sheel, Maarit A. Laaksonen, Lin Zhu, Yiyi Lin, Christina Abdel Shaheed, Mouna Sawan, Assel Mussagulova, Danijela Gnjidic, Pani Patu, Bette Liu, Fiona F Stanaway

## Abstract

**Background:** International evidence has documented ethnic inequalities in COVID-19 vaccine uptake, but national evidence for Australia remains limited. We aimed to quantify ethnic inequalities in COVID-19 vaccine uptake in the first 6 months of 2024 and examine retrospective trends in Dose 1–4 (2021–22) across detailed ethnic groups among older adults.

**Methods:** We conducted a nationwide cohort study of Australian residents aged ≥75 years who were not Aboriginal or Torres Strait Islander (N=2,038,522) by linking the 2021 Census, Australian Immunisation Register, death, and migration data. Age-standardized uptake of any COVID-19 vaccine dose by ethnic group was calculated (Jan 1–June 30, 2024). Stratified descriptive analyses were conducted to explore intersections between ethnicity and other key sociodemographic characteristics. Uptake of Dose 1–4 during 2021–22 was also assessed across ethnic groups.

**Results:** In the first 6 months of 2024, 31.1% of the cohort received a COVID-19 vaccine. Uptake was substantially lower in several ethnic groups, including Central Asian (<10.0%, 95% CI <10.0–10.7), North African and Middle Eastern (<10.0%, 95% CI <10.0–<10.0), Pasifika (13.0%, 95% CI 11.7–14.4), and South Eastern European (10.5%, 95% CI 10.3–10.7) groups. These differences persisted even among individuals born in Australia, with higher English proficiency, higher educational attainment, and living in less disadvantaged areas. Similar inequalities were observed across earlier vaccine doses.

**Conclusions:** Substantial ethnic inequalities in COVID-19 vaccination uptake persist among older Australians. Reliance on country of birth, language, or socioeconomic factors alone does not fully identify groups with the lowest uptake. Incorporating more detailed ethnicity information may improve identification of under-served groups and inform more targeted and culturally appropriate vaccination strategies.

## Introduction

Australia achieved high COVID-19 vaccination coverage during the initial rollout, with over 95% of individuals aged ≥16 years receiving two doses by the end of 2022.^1^ However, uptake of recommended booster doses among older adults has declined substantially. Despite the Australian Technical Advisory Group on Immunisation (ATAGI) recommending a 6-monthly vaccination interval for adults aged ≥75 years,^2^ only 23.2% received a dose between July and December 2023.^3^ Older adults remain at highest risk of severe outcomes, accounting for more than 80% of COVID-19 deaths in 2023.^4^ Those who had not received a vaccine dose within the previous 6 months experienced a two-fold higher risk of COVID-19-related death.^5^

Ethnic inequalities in COVID-19 vaccination uptake have been reported in several high-income countries where ethnicity is systematically measured in national linked administrative or large-scale survey datasets.^6–8^ In Australia, however, publicly available data remains limited because ethnicity is not routinely collected in administrative health dataset.^9^ Previous Australian studies have examined vaccination inequalities using country of birth, English proficiency, and other sociodemographic indicators.^10,11^

While these measures have informed vaccination strategies, it remains unclear whether they adequately identify all groups with low uptake. Country of birth and language-related measures may not capture the full diversity of Australia’s population, and broad categories may mask important differences between smaller ethnic groups. This limitation is particularly important among older adults, for whom booster vaccination remains critical for protection against severe COVID-19 outcomes.

In this study, we examined ethnic inequalities in COVID-19 vaccine uptake among Australians aged ≥75 years from 01 January to 30 June 2024 using detailed ethnic group classifications derived from linked national datasets. We also assessed whether commonly used indicators captured these differences and examined uptake of Dose 1–4 during 2021–22 to provide historical context.

## Methods

### Data Sources and Linkage

The Person-Level Integrated Data Asset (PLIDA) is a secure data resource managed by the Australian Bureau of Statistics (ABS) that links national Census data with administrative health and social datasets.^12^ For this study, 2021 Census data were linked with COVID-19 vaccination records from the Australian Immunisation Register (AIR), death registrations, and the Migration dataset via the Person Linkage Spine Version 6 (the Spine). The Spine is built from the Medicare Consumer Directory, Data Over Multiple Individual Occurrences (DOMINO) Centrelink Administrative Data, and Personal Income Tax datasets, and enables linkage across multiple datasets,^13^ with a high linkage rate of 96.3% for the 2021 Census.

Australia conducts a compulsory national Census every five years, with the most recent in August 2021 achieving a 96.1% response rate for private dwellings.^14^ The Census provides demographic and socioeconomic information. The AIR records all vaccines administered in Australia.^15^ Death registrations and the Migration dataset capture all deaths and monthly absences from Australia respectively, up to 31 December 2023.

### Study Population

The primary study population comprised non-Indigenous Australian residents aged ≥75 years as of 2024 who responded to the 2021 Census. This age threshold aligns with ATAGI’s recommendation, issued in September 2023, for 6-monthly vaccination among people aged ≥75 years.^2^ Individuals were excluded if they: 1) were identified as visitors in the Census, 2) could not be linked to the Spine, 3) had missing or inadequately described ethnicity, or 4) had died or left Australia prior to the study commencement on 01 January 2024.

The study population was limited to non-Indigenous Australians, consistent with consultation and the expressed preference of Aboriginal and Torres Strait Islander peoples to avoid direct comparisons with other ethnic groups.

To provide historical context on COVID-19 vaccine equality, secondary analysis was conducted on a separate 2021 cohort during the pandemic (see Section 1.1 of Supplementary Material for details).

### Exposure and Key Characteristics

The analysis included ethnicity as the primary exposure, alongside age, sex, country of birth, English proficiency, remoteness area, educational attainment, nursing home residence, socioeconomic position, and self-reported long-term health conditions.

Ethnicity was based on ancestry responses in the 2021 Census and classified using the Australian Standard Classification of Cultural and Ethnic Groups (ASCCEG),^16^ following the approach described by Stanaway et al.^17^ For primary analysis, ethnic groups were categorized into 8 broad groups (14 subgroups): Asian (Central Asian, Other Asian), European (Eastern European, South Eastern European, Southern European, Other European), Māori, Multiethnic, North African and Middle Eastern (Jewish, Other North African and Middle Eastern), North and Latin American (North American, Other North and Latin American), Pasifika, and Sub-Saharan African. Subgroups were based on important differences in vaccination rates between either individual ethnic groups (e.g. Jewish) or several ethnic groups in a small region (e.g Central Asian) and the larger continent level grouping (e.g. Asian). See Section 1.1 of the Supplementary Materials for details.

Based on literature review and expert consultation,^10,11,18^ demographic characteristics were also derived from the Census and included age, sex (male, female), country of birth (born in Australia, born overseas), and English proficiency (very well/ well, not well/ not at all). Remoteness area was classified into two categories (major cities, non-major cities).^19^ Nursing home residence (yes/no) was identified based on the ‘Type of non-private dwelling’ variable. Socioeconomic position was assessed using educational attainment (postgraduate/bachelor’s degree, no bachelor’s degree) and area-level disadvantage. Area-level disadvantage was measured using the Socio-Economic Indexes for Areas (SEIFA) Index of Relative Socio-economic Disadvantage (IRSD) based on Statistical Area Level 2 (SA2) and grouped into three categories (less disadvantage, moderate disadvantage, greater disadvantage).^20^ Self-reported long-term health condition status was categorized into no long-term health condition or one or more long-term health conditions.

### Outcome

The primary outcome was age-standardized uptake of any COVID-19 vaccine dose, defined as the percentage receiving at least one COVID-19 vaccine dose between 01 January and 30 June 2024. This period reflects the implementation of the 6-monthly booster recommendation for older adults.^2^ Secondary outcomes included the age-standardized uptake of Dose 1–4 from 10 August 2021 to 31 December 2022 (details provided in Section 1.1 of Supplementary Material).

### Statistical Analyses

All descriptive analyses were age-standardized using the World Health Organization (WHO) standard population (2000–25).^21^ All counts or proportions under 10 were suppressed and all COVID-19 vaccination results reporting counts or rates were rounded probabilistically to the nearest multiple of five according to the confidentiality guidelines of ABS to minimise disclosure risk. All analyses were performed in R (version 4.4.1).

For the primary analysis, age-standardized uptake of COVID-19 vaccine dose by ethnic group was calculated for the period 01 January to 30 June 2024 (inclusive). Migration/traveller’s and death registry data were used to define the study population at the start of the study. Stratified descriptive analyses were conducted to explore intersections between ethnicity and other key sociodemographic characteristics.

Sensitivity analyses extended the observation period to 30 September 2024 (9 months) to minimize potential misclassification related to temporary vaccine deferral. For covariates with >5% missing data, extreme-case analyses were conducted by separately recoding missing values into each category of the covariate.

For the secondary analysis involving the 2021 cohort, age-standardized COVID-19 vaccination uptake for Dose 1–4 was estimated for each ethnic group from 10 August 2021 to 31 December 2022 (details provided in Section 1.2 of Supplementary Material).

### Ethics approval

The analysis of unit record data with a waiver of consent was approved by the New South Wales Population and Health Services Research Ethics Committee (2020/ETH01066).

## Results

A total of 2,038,522 individuals (median [IQR (interquartile range)] age, 77 [74–82] years) were included in the primary analysis. The sociodemographic characteristics of the study population by ethnic group are shown in Table 1. Study population exclusion flowchart is shown in Figure S1.

**Table 1.**
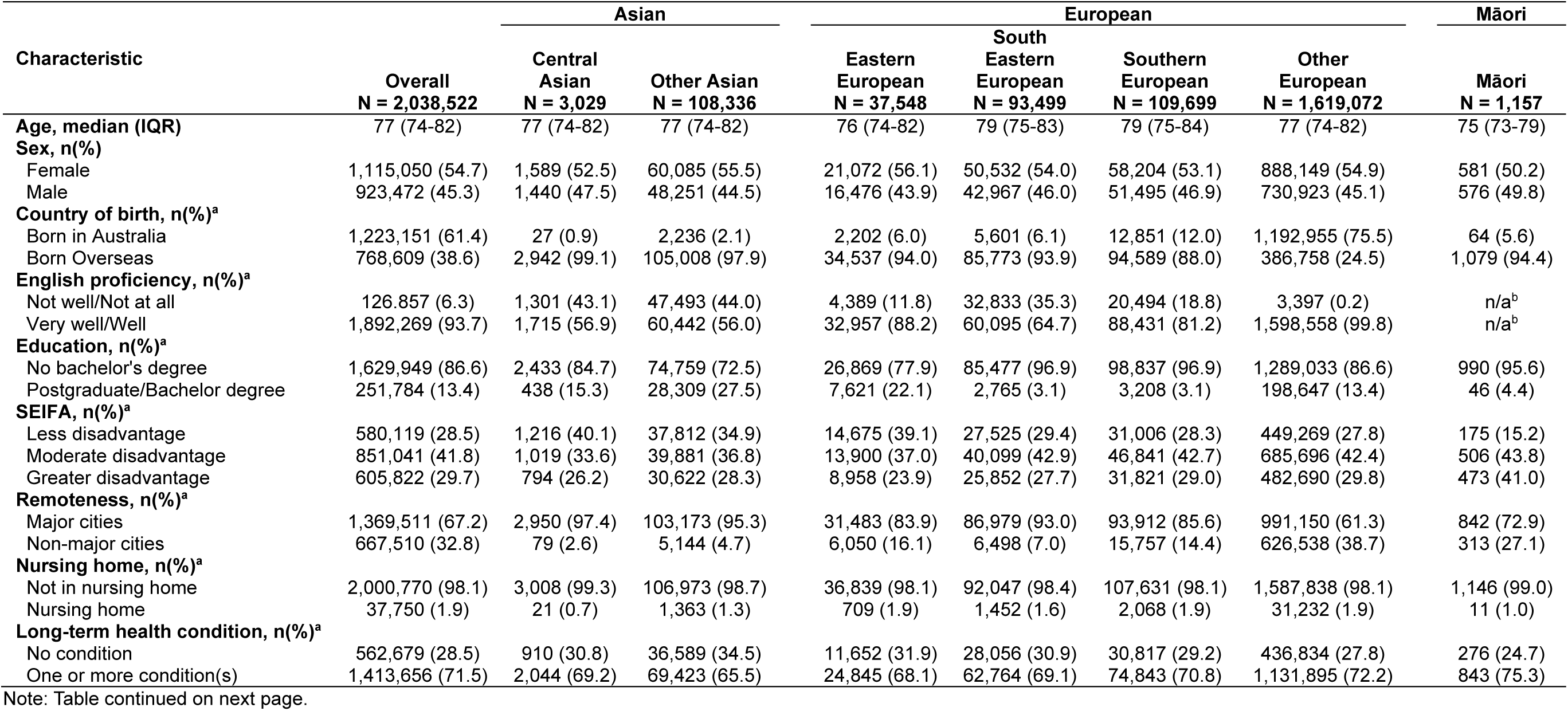
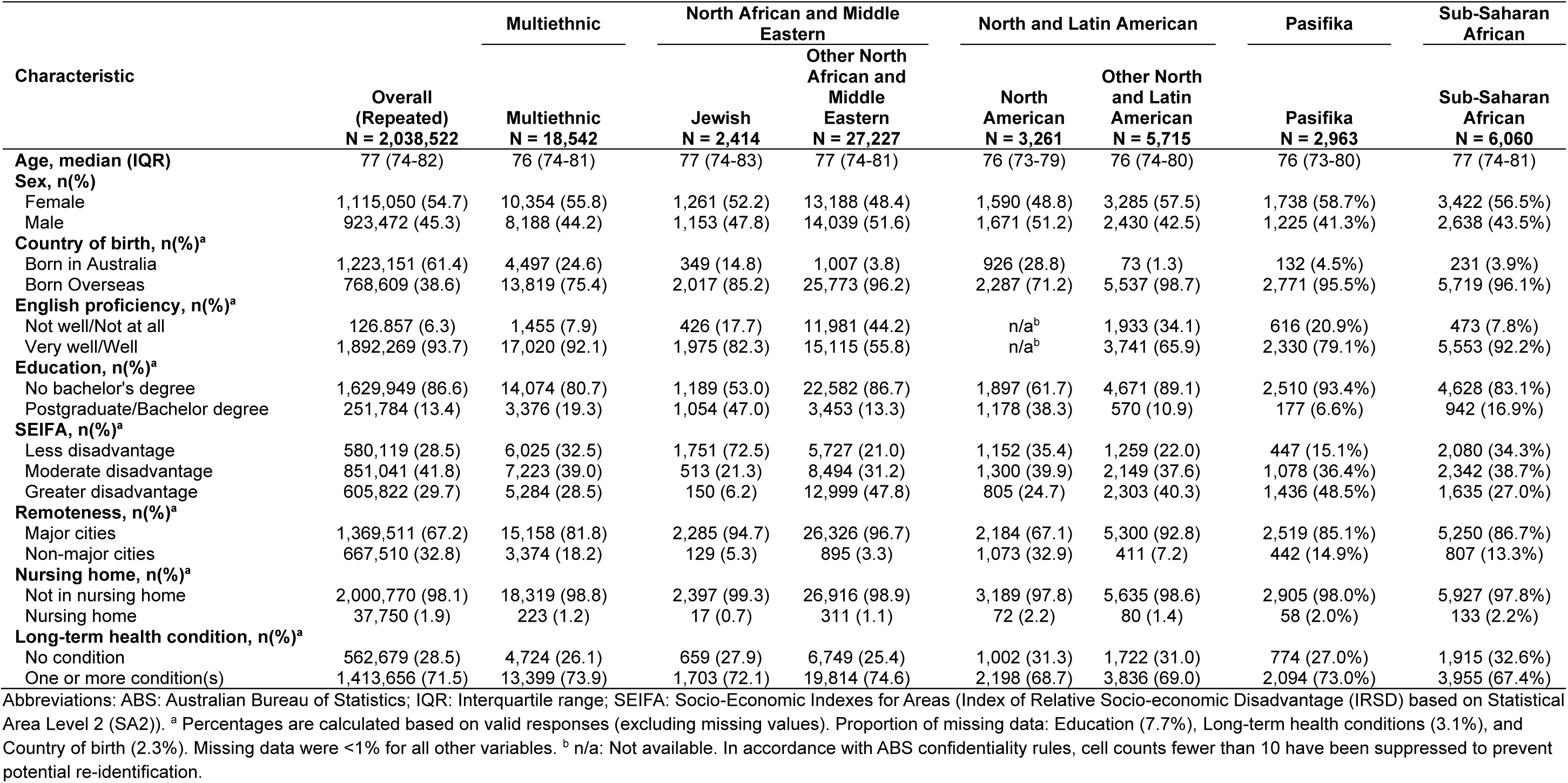
Characteristics of the Study Population by Ethnic Groups at Baseline for Cohort Aged 75+ years as of 2024.

### COVID-19 Vaccine Dose Uptake by Ethnicity (Jan–Jun 2024)

During the first 6 months of 2024, the overall age-standardized rate for receiving at least one COVID-19 vaccine dose was 31.1%. Rates were low among several broad ethnic groups, including Asian (16.9%, 95%CI: 16.7–17.1%; with <10.0% [<10.0–10.7%] for the Central Asian subgroup), North African and Middle Eastern (<10.0%, <10.0–<10.0%), and Pasifika (13.0%, 11.7–14.4%). While the broader European group had a higher rate (32.6%, 32.5–32.7%), only 10.5% (10.3–10.7%) of South Eastern Europeans received a dose. In contrast, uptake among Jewish (27.0%, 24.9–29.3%) and North American (35.3%, 33.3–37.5%) groups was substantially higher than that observed in their respective broad groups of North African and Middle Eastern and North and Latin American (Figure 1). Results were similar when the analysis was extended to the nine-month period (01 January–30 September 2024) to account for delayed vaccination due to recent SARS-CoV-2 infection, with an overall age-standardized rate of 37.6% (Figure S2).

**Figure 1.**
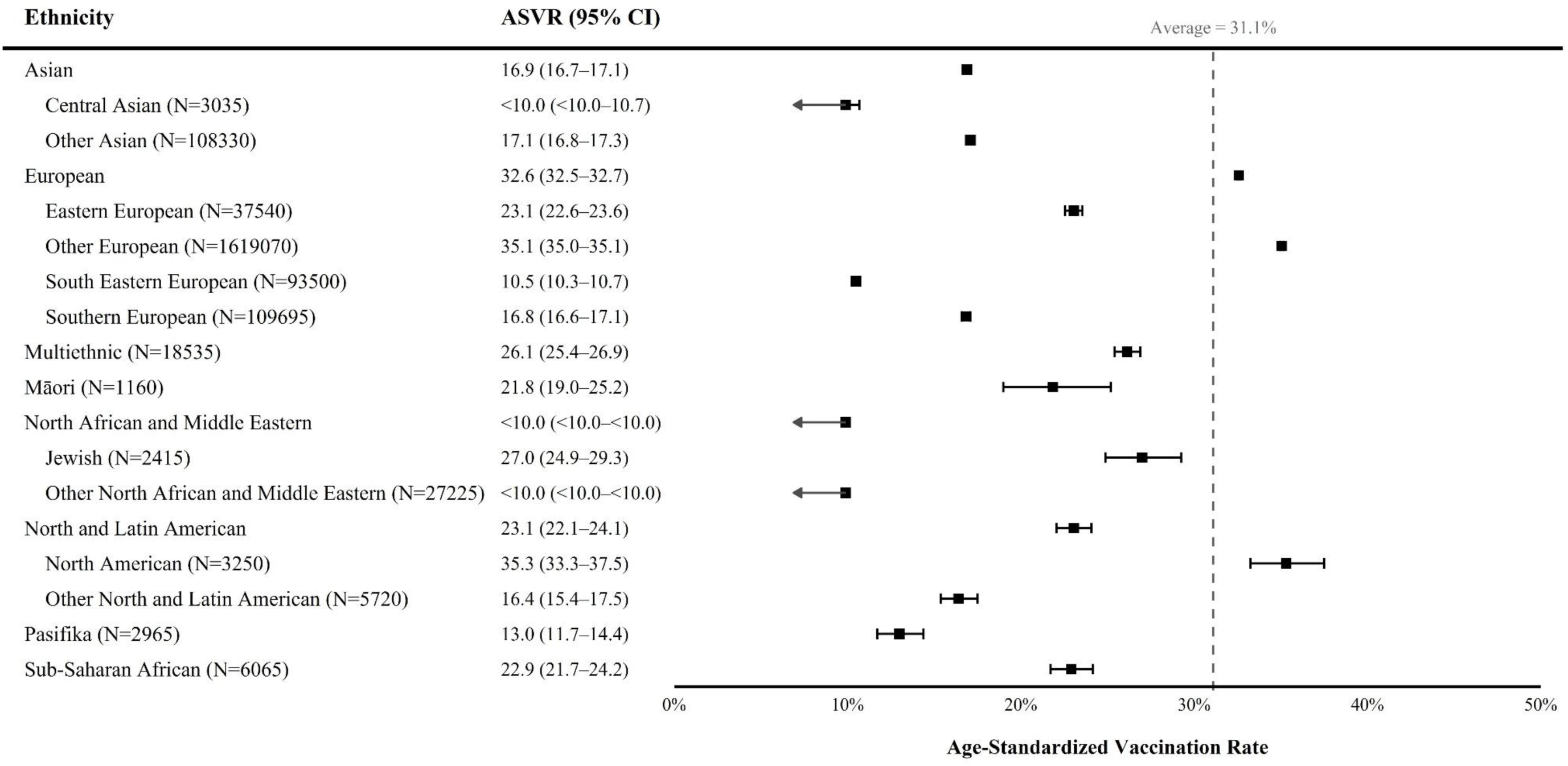
Age-standardized Uptake of COVID-19 Vaccine Dose by Ethnicity, Jan 1–June 30, 2024. Note. In accordance with Australian Bureau of Statistics confidentiality rules, percentages below 10% were displayed as “<10.0”. COVID-19 vaccination data were probabilistically rounded to the nearest multiple of five. Rates are age-standardized to the World Health Organization standard population. N: number of individuals in this ethnic group, with counts probabilistically rounded to the nearest multiple of five; ASVR: Age-standardized vaccination rate (%); 95% CI: 95% Confidence Interval.

### Interaction of COVID-19 Vaccine Dose Uptake by Ethnicity and Key Characteristics (Jan–Jun 2024)

Overall, age-standardized uptake of a COVID-19 vaccine was higher among individuals with higher English proficiency (32.6% vs <10%), born in Australia (35.7% vs 24.1%), with higher education (39.9% vs 30.0%), with long-term health conditions (32.3% vs 28.7%), and living in less disadvantaged areas (36.1% vs 26.7%), non-major cities (32.3% vs 30.5%), and nursing homes (38.0% vs 31.0%). Nevertheless, important ethnic differences persisted within these subgroups, even among those with characteristics associated with higher vaccine uptake (Figure 2–3, Figure S3–7). No differences were observed by sex (31.3% in males vs 30.9% in females) (Figure S8). Result of sensitivity analyses for variables with >5% missingness (e.g., education) were consistent with the main results.

**Figure 2.**
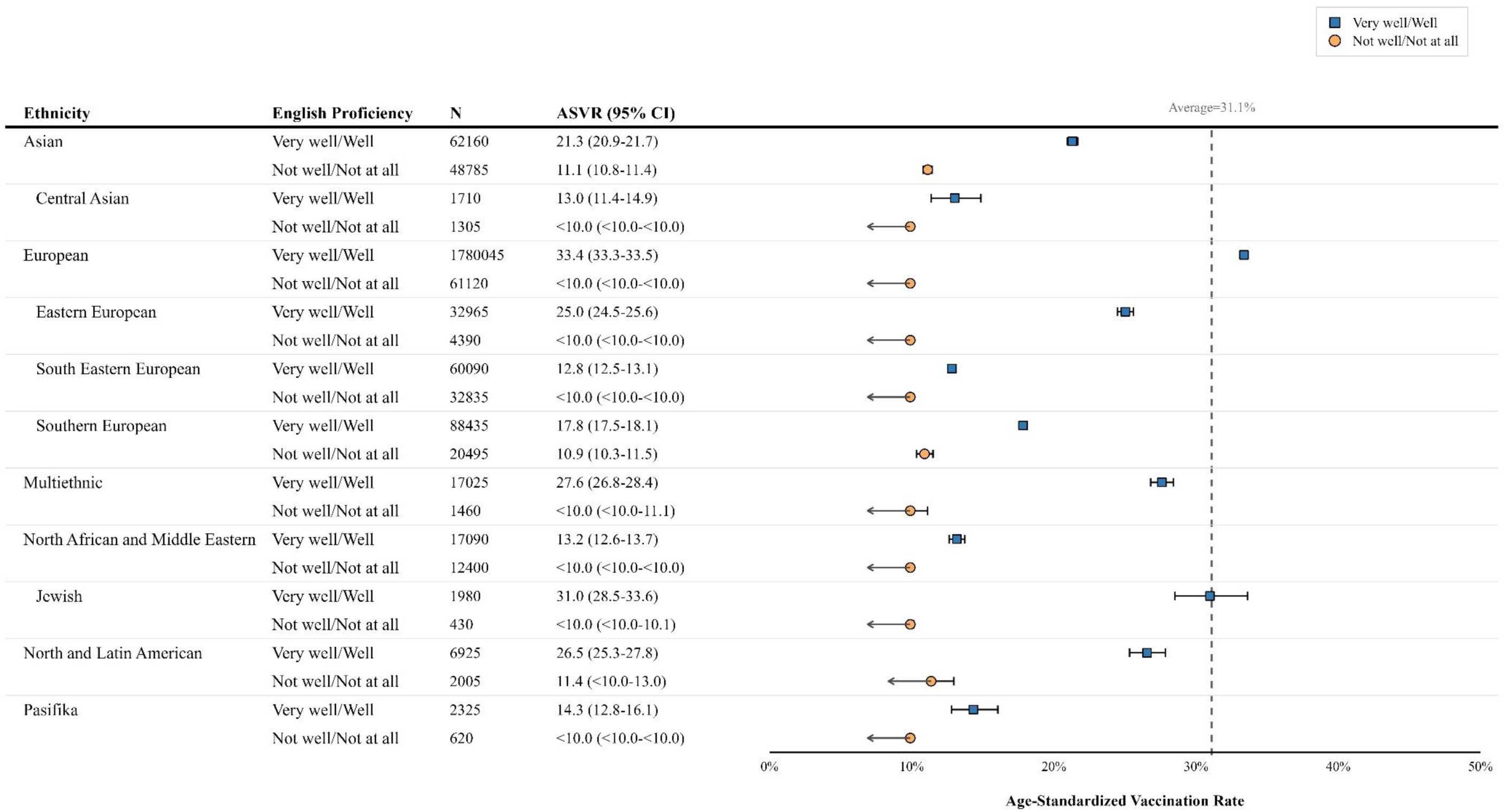
Age-standardized Uptake of COVID-19 Vaccine Dose by Ethnicity and English Proficiency, Jan 1–June 30, 2024 Note. In accordance with ABS confidentiality rules, cell counts fewer than 10 were suppressed to prevent potential re-identification; therefore, the Māori and North America group are not shown in this figure. Percentages below 10% were displayed as “<10.0”. COVID-19 vaccination data were probabilistically rounded to the nearest multiple of five. Rates are age-standardized to the World Health Organization standard population. N: number of individuals in this ethnic group, with counts probabilistically rounded to the nearest multiple of five; ASVR: Age-standardized vaccination rate (%); 95% CI: 95% Confidence Interval.

**Figure 3.**
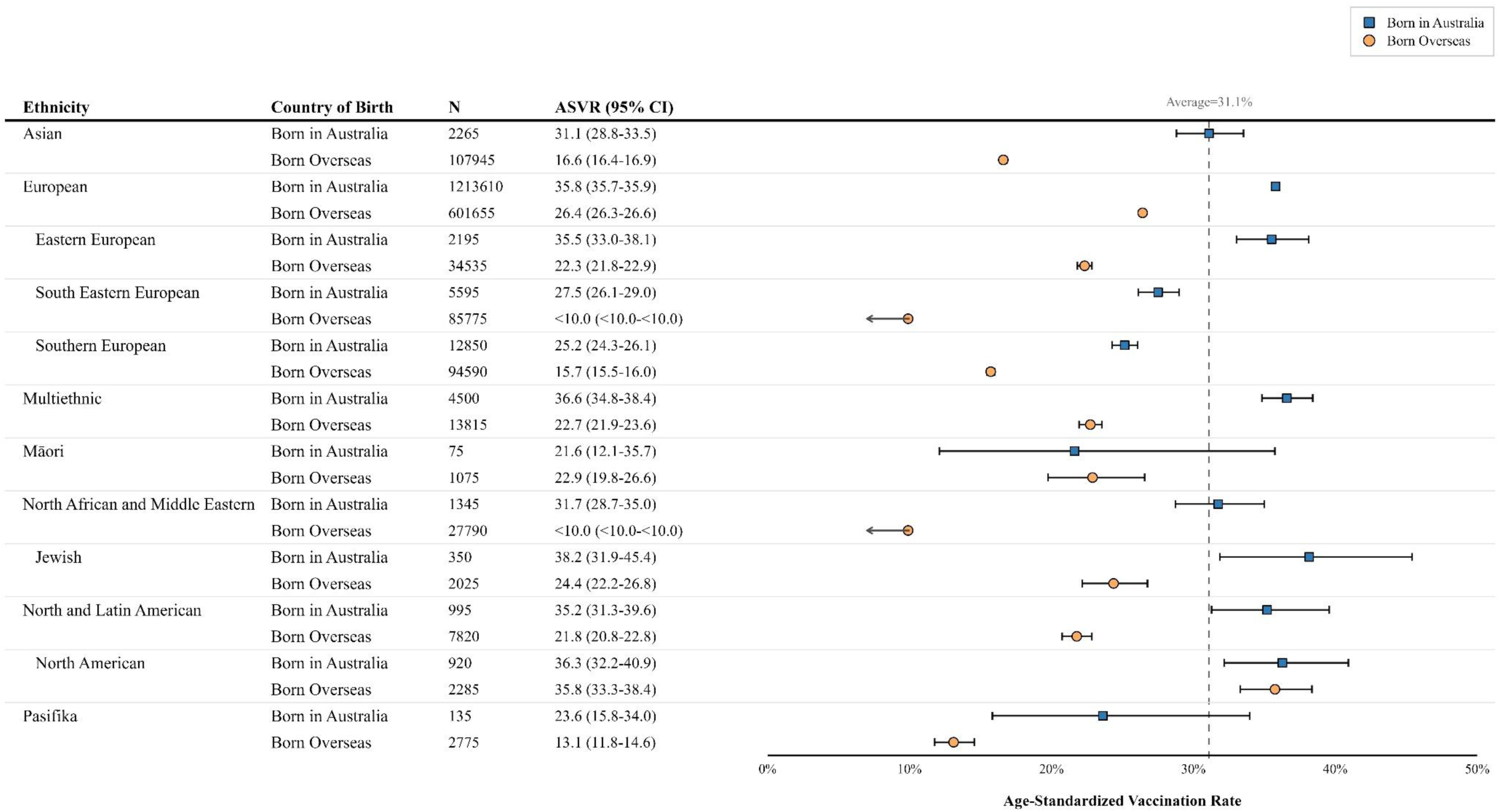
Age-standardized Uptake of COVID-19 Vaccine Dose by Ethnicity and Country of Birth, Jan 1–June 30, 2024 Note. In accordance with Australian Bureau of Statistics confidentiality rules, cell counts fewer than 10 were suppressed to prevent potential re-identification; therefore, the Central Asian group is not shown in this figure. Percentages below 10% were displayed as “<10.0”. COVID-19 vaccination data were probabilistically rounded to the nearest multiple of five. Rates are age-standardized to the World Health Organization standard population. N: number of individuals in this ethnic group, with counts probabilistically rounded to the nearest multiple of five; ASVR: Age-standardized vaccination rate (%); 95% CI: 95% Confidence Interval.

Among those with self-rated English proficiency of not well or not at all, there was little difference in uptake by ethnicity, with uptake below 11.5% across all ethnic groups (Figure 2). However, even among those who reported speaking English well/very well, there were important differences by ethnicity, with uptake below 15% in Central Asian (13.0%, 95%CI: 11.4–14.9%), North African and Middle Eastern (13.2%, 12.6–13.7%), Pasifika (14.2%, 12.8–16.1%), and South Eastern European (12.8%, 12.5–13.1%) groups.

Ethnic inequalities were observed among people born overseas, with lower uptake observed in Asian (16.6%, 16.4–16.9%), North African and Middle Eastern (<10.0%, <10.0–<10.0%), Pasifika (13.1%, 11.8–14.6%), South Eastern European (<10.0%, <10.0–<10.0%), and Southern European (15.7%, 15.5–16.0%) groups (Figure 3). Although uptake was higher among those born in Australia than among those born overseas in all ethnic groups except for Māori communities, the size of this difference varied by ethnicity and inequalities remained. The largest difference was seen in the North African and Middle Eastern group, rising from below 10% among those born overseas to 31.7% among those born in Australia. The smallest difference was observed in the North American group (35.8% vs 36.3%). Within the Australian-born population, uptake remained relatively low in the Māori, Pasifika, South Eastern European, and Southern European groups.

A similar pattern was observed for socioeconomic indicators. Uptake among Central Asian (14.0%, 10.7–18.1%) and North African and Middle Eastern (18.9%, 17.7–20.3%) groups remained low even among individuals with a postgraduate/ bachelor’s degree (Figure S3). Likewise, uptake in the Central Asian (14.6%, 12.5–16.9%), North African and Middle Eastern (17.5%, 16.6–18.5%), and South-Eastern European (13.7%, 13.3–14.2%) groups remained comparatively low among those living in areas of less disadvantage (Figure S5).

### Patterns of COVID-19 Vaccine Uptake for Dose 1–4 (August 2021–December 2022)

In the secondary analysis, 1,671,996 individuals aged ≥75 years were included (median age 80 years [IQR 77–85]). Sociodemographic characteristics, study population exclusion flowchart and the study period for each dose are shown in Table S1 and Figure S9.

By 31 December 2022, overall vaccination coverage reached 96% for both the first and second doses, 91% for the third dose, and 74% for the fourth dose. Across all doses, coverage was slightly higher among males than females (Figure S10).

On 10 August 2021, the lowest first- and second-dose coverage was observed among Central Asian (57%; 22%), North African and Middle Eastern (54%; 27%), Pasifika (59%; 30%), and South Eastern European (54%; 23%) groups (Figure S10–13). However, by the end of 2021, differences in first- and second-dose coverage between ethnic groups had diminished, with overall coverage reaching high levels. For the third and fourth doses, lower coverage persisted among the same four ethnic groups throughout the study period. By the end of 2022, coverage remained lowest among Central Asian (72% for the third dose; 38% for the fourth dose), North African and Middle Eastern (66%; 37%), Pasifika (75%; 44%), and South Eastern European (73%; 41%) groups. Interactive versions of Figures S11–14, allowing users to select and compare specific ethnic groups, are available in the OSF repository: https://osf.io/wgtpk/files.

## Discussion

In this national linked-data study of Australians aged ≥75 years, we observed ethnic inequalities in uptake of the COVID-19 vaccine during the first half of 2024. Overall uptake was low, with low rates observed particularly among Central Asian, North African and Middle Eastern, Pasifika, and South Eastern European groups. These differences persisted even among individuals born in Australia, with higher English proficiency, higher educational attainment, and living in less disadvantaged areas. Historical uptake patterns for Dose 1–4 during 2021–22 showed a similar pattern: early delays in uptake for the first and second doses largely diminished by the end of 2021, but inequalities reappeared and widened for the third and fourth doses, concentrated in the same ethnic groups as observed in the first half of 2024.

Australia’s COVID-19 Vaccine Rollout Strategy initially achieved high primary uptake among older adults through phased eligibility and targeted outreach,^22^ which minimized differences between ethnic groups. As we have transitioned out of the emergency phase of the pandemic, COVID-19 vaccine uptake among older adults in 2024 has decreased and annual coverage is approximately half that of influenza vaccination in Australia.^23^ This suggests that factors beyond convenience and accessibility, such as perceived risk and vaccine fatigue,^24^ may influence COVID-19 vaccine uptake and could vary across ethnic groups.

Our findings show that vaccination inequalities in Australia were concentrated in Central Asian, North African and Middle Eastern, Pasifika, and South Eastern European groups. These patterns are partly consistent with previous Australian evidence. For example, the Grattan Report identified lower uptake among people born in the Middle Eastern and South Eastern Europe.^11^ However, reliance on country-of-birth classifications can obscure important within-group differences. Māori and Pasifika populations, for example, are often grouped within a broad “Oceania” category that includes all those born in New Zealand, many of whom are of European ethnicity. Country of birth does not capture those born in Australia who identify as ethnically diverse, nor does it distinguish between people born overseas whose ethnicity is not indicated by their country of birth. Similarly, smaller population groups such as Central Asians, who had particularly low uptake in our study, are usually combined with other larger groups due to limited sample sizes. These approaches can mask substantial heterogeneity in vaccination uptake and limit the ability to identify specific groups with particularly low uptake.

Consistent with previous studies,^6,10,11,25–27^ lower uptake was observed among individuals born overseas, with lower education levels, residing in areas of greater socioeconomic disadvantage, or with limited English proficiency. However, these factors may still obscure important within-group differences and fail to fully explain persistent inequalities. For example, for ethnic groups with consistently low vaccination uptake, including North African and Middle Eastern, Central Asian, South Eastern European, and Pasifika populations, uptake remained below 15% even among individuals reporting that they spoke English “very well” or “well”. This indicates that multilingual translation of health messaging is necessary but insufficient to fully eliminate the structural and contextual barriers that can impact vaccination in these groups.^28^

Beyond language, non-majority ethnic groups face additional barriers shaped by differences in access to health information and prior lived experiences. These include variations in health literacy, patterns of engagement with mainstream digital platforms, and experiences of discrimination, all of which can undermine trust in mainstream health messaging and increase exposure to misinformation.^28–30^ Vaccine decisions are also strongly influenced by trusted informal networks such as family and friends, community or religious leaders, and media from countries of origin, as well as culturally shaped norms and beliefs, rather than mainstream sources.^31^ For example, a recent study found low vaccine confidence in South Eastern Europe (specifically North Macedonia and Bosnia and Herzegovina) and Central Asia,^32^ which may be reflected among corresponding migrant populations in Australia. These individual, social, and structural influences may help explain the persistent ethnic inequalities identified in our study.

## Limitations

Due to the unavailability of data on notifiable COVID-19 cases, we could not account for SARS-CoV-2 infection status over time across ethnic groups, which may have influenced subsequent vaccination decisions. However, our sensitivity analysis extending the study window to nine months demonstrated that inequalities in uptake persisted. Furthermore, analyses were restricted to COVID-19 vaccination among older adults and may not generalise to other age groups or vaccination programmes. Finally, key determinants of vaccine uptake, including behavioural, attitudinal, and health service access factors,^29,31,33^ were not captured in national administrative data, limiting our ability to directly examine some of the mechanisms underlying the observed ethnic inequalities.

Monitoring ethnic inequalities in vaccination uptake is essential for equitable public health systems in multicultural societies. Current approaches in Australia often rely on indicators such as country of birth, language, or socioeconomic factors, which may not fully capture groups with low uptake. Consistent with recent Australian reviews,^18,34^ our findings highlight the importance of routinely collecting and using detailed ethnicity data to better identify under-served groups, including within populations already recognised as being at higher risk of severe COVID-19 outcomes. Such information can support more targeted vaccination strategies that are responsive to the needs of specific communities, including culturally grounded and trust-based approaches involving community leaders and diverse healthcare workers, rather than reliance on translated health information alone.^28,31,35^

## Conclusions

Substantial ethnic inequalities in COVID-19 vaccine uptake were observed among Australians aged ≥75 years. These differences persisted even among individuals with characteristics typically associated with higher uptake. Reliance on country of birth, language, or socioeconomic factors alone did not fully identify groups with the lowest uptake. Incorporating detailed ethnicity information may improve identification of under-served groups and inform more targeted and culturally appropriate vaccination strategies in Australia.

## Data Availability

The data used in this study are available through the Person-Level Integrated Data Asset (PLIDA), managed by the Australian Bureau of Statistics. Access to these data is subject to approval and cannot be shared publicly. Researchers may apply for access through the Australian Bureau of Statistics following the relevant governance and ethical approval processes.

