## Supplementary material for "Ethnic Inequalities in COVID-19 Vaccination Uptake Among Older Adults in Australia: A Nationwide Linked Data Study"

Peiyao Xu

### Section 1. Supplementary Methods for the 2021 Cohort

##### 1.1 Identification and Definition of the 2021 Cohort

**Data Sources and Linkage**

Consistent with the primary analysis, this study utilized data from the 2021 Australian Census data, linked with the Australian Immunisation Register (AIR), death registrations, and the Migration dataset via the Person Linkage Spine Version 6 (the Spine) (details provided in the Methods section of the main text).

**Study Population**

The study population comprised non-Indigenous Australian residents aged ≥75 years as of 2021 who responded to the 2021 Census. Individuals were excluded if they: 1) were identified as visitors in the Census, 2) could not be linked to the Spine, 3) had missing or inadequately described ethnicity.

**Ethnicity Classification**

Ethnicity was based on ancestry responses in the Census. Classification of ethnic groups followed the approach described by Stanaway et al.,[1] which prioritises ethnic minority identities over national identities, groups individuals reporting multiple ancestries into multiethnic categories, and maintains the highest level of granularity where meaningful health inequalities exist. We used the Australian Standard Classification of Cultural and Ethnic Groups (ASCCEG) to guide this definition,[2] starting from "Cultural and ethnic group" (the most detailed level; small regional areas), then aggregating to "Narrow group" (larger regional areas), and finally to "Broad group" (continents), collapsing categories only when no meaningful differences were observed. This was based on consistency of point estimates and overlap of confidence intervals, similar to the process of assessing heterogeneity in Grading of Recommendations Assessment, Development and Evaluation (GRADE).[3] This classification approach balances granularity of categories with the need for better precision, minimisation of disclosure risks, and ease of interpretability.

For this secondary analysis, to ensure statistical stability and simplify reporting, ethnic groups were categorized into 8 broad groups (13 subgroups): Asian (Central Asian, Other Asian), European (Eastern European, South Eastern European, Other European), Māori, Multiethnic, North African and Middle Eastern (Jewish, Other North African and Middle Eastern), North and Latin American (North American, Other North and Latin American), Pasifika, and Sub-Saharan African. The 'Other European' category mainly comprises Anglo-Celtic and Northern and Western European ancestries, representing the largest ethnic group in Australia.

**Outcome**

The secondary outcomes were age-standardized COVID-19 vaccination uptake of Dose 1, 2, 3, and 4 from the 2021 Census night (10 August 2021) to 31 December 2022.

##### 1.2 Statistical Analyses of the 2021 Cohort

Age-standardized COVID-19 vaccination uptake for the first and second doses was estimated for each ethnic group from 10 August 2021 to 31 December 2022. In Australia, recommendation for the administration of third doses to the non-immunocompromised population commenced on 11 October 2021,[4] followed by fourth doses for all adults aged 65+ years from 25 March 2022.[5] To avoid disclosure of small counts, third- and fourth-dose rates were calculated for the periods 31 December 2021 to 31 December 2022 and 30 June 2022 to 31 December 2022, respectively. Migration/traveller's and death registry data were used to dynamically update the denominator by removing individuals who exited the country or passed away during the follow-up period.

#### Section 2. Supplementary Tables

##### Table S1. Characteristics of the Study Population by Ethnic Groups at Baseline for the 2021 cohort (Part 1 of 2)

| Characteristic | Overall | Asian | | European | | | Māori | Multiethnic |
|---|---|---|---|---|---|---|---|---|
| | Overall N = 1,671,996 | Central Asian N = 2,441 | Other Asian N = 84,422 | Eastern European N = 29,132 | South Eastern European N = 83,307 | Other European N = 1,420,731 | Māori N = 803 | Multiethnic N = 13,933 |
| **Age, median (IQR)** | 80 (77-85) | 80 (77-85) | 80 (77-85) | 81 (77-86) | 81 (78-85) | 80 (77-85) | 78 (76-81) | 80 (77-84) |
| **Sex, n(%)** | | | | | | | | |
| Female | 920,562 (55.1) | 1,299 (53.2) | 46,696 (55.3) | 16,519 (56.7) | 44,196 (53.1) | 784,515 (55.2) | 422 (52.6) | 7,819 (56.1) |
| Male | 751,434 (44.9) | 1,142 (46.8) | 37,726 (44.7) | 12,613 (43.3) | 39,111 (46.9) | 636,216 (44.8) | 381 (47.4) | 6,114 (43.9) |
| **Country of birth, n(%)[a]** | | | | | | | | |
| Born in Australia | 998,596 (61.3) | 24 (1.0) | 1,810 (2.2) | 1,673 (5.9) | 4,768 (5.9) | 984,684 (71.2) | 50 (6.3) | 3,387 (24.7) |
| Born Overseas | 631,735 (38.7) | 2,376 (99.0) | 81,716 (97.8) | 26,793 (94.1) | 76,689 (94.1) | 398,694 (28.8) | 744 (93.7) | 10,353 (75.3) |
| **English proficiency, n(%)[a]** | | | | | | | | |
| Not well/Not at all | 121,888 (7.4) | 1,104 (45.4) | 41,556 (49.5) | 4,455 (15.4) | 32,790 (39.6) | 26,841 (1.9) | n/a[b] | 1,338 (9.6) |
| Very well/Well | 1,530,648 (92.6) | 1,326 (54.6) | 42,436 (50.5) | 24,461 (84.6) | 49,910 (60.4) | 1,376,006 (98.1) | n/a[b] | 12,535 (90.4) |
| **Education, n(%)[a]** | | | | | | | | |
| No bachelor's degree | 1,338,548 (88.1) | 1,993 (85.9) | 59,024 (73.9) | 21,067 (80.2) | 76,201 (97.5) | 1,139,278 (88.7) | 665 (96.0) | 10,775 (82.5) |
| Postgraduate/Bachelor degree | 181,538 (11.9) | 326 (14.1) | 20,829 (26.1) | 5,202 (19.8) | 1,963 (2.5) | 145,636 (11.3) | 28 (4.0) | 2,288 (17.5) |
| **SEIFA, n(%)[a]** | | | | | | | | |
| Less disadvantage | 472,500 (28.3) | 1,024 (42.0) | 29,195 (34.6) | 11,300 (38.8) | 24,965 (30.0) | 391,832 (27.6) | 125 (15.6) | 4,427 (31.8) |
| Moderate disadvantage | 696,098 (41.7) | 799 (32.7) | 31,059 (36.8) | 10,671 (36.6) | 35,754 (42.9) | 599,707 (42.2) | 341 (42.6) | 5,456 (39.2) |
| Greater disadvantage | 502,491 (30.1) | 618 (25.3) | 24,153 (28.6) | 7,154 (24.6) | 22,573 (27.1) | 428,345 (30.2) | 335 (41.8) | 4,042 (29.0) |
| **Remoteness, n(%)[a]** | | | | | | | | |
| Major cities | 1,134,399 (67.9) | 2,380 (97.5) | 80,690 (95.6) | 24,799 (85.1) | 77,716 (93.3) | 902,197 (63.5) | 598 (74.6) | 11,534 (82.8) |
| Non-major cities | 536,718 (32.1) | 61 (2.5) | 3,720 (4.4) | 4,326 (14.9) | 5,577 (6.7) | 517,710 (36.5) | 204 (25.4) | 2,391 (17.2) |
| **Nursing home, n(%)[a]** | | | | | | | | |
| Not in nursing home | 1,596,895 (95.5) | 2,399 (98.3) | 82,089 (97.2) | 27,685 (95.0) | 80,430 (96.5) | 1,353,890 (95.3) | 789 (98.3) | 13,552 (97.3) |
| Nursing home | 75,099 (4.5) | 42 (1.7) | 2,333 (2.8) | 1,447 (5.0) | 2,877 (3.5) | 66,839 (4.7) | 14 (1.7) | 381 (2.7) |
| **Long-term health condition, n(%)[a]** | | | | | | | | |
| No condition | 394,795 (24.4) | 660 (27.7) | 24,401 (29.5) | 7,481 (26.4) | 21,613 (26.7) | 328,815 (23.9) | 175 (22.4) | 3,026 (22.1) |
| One or more condition(s) | 1,225,941 (75.6) | 1,725 (72.3) | 58,303 (70.5) | 20,837 (73.6) | 59,453 (73.3) | 1,046,713 (76.1) | 605 (77.6) | 10,636 (77.9) |

Note: Table continued on next page.

**Table S1 (Continued). Characteristics of the Study Population by Ethnic Groups at Baseline for the 2021 cohort (Part 2 of 2)**

| Characteristic | Overall (Repeated) N = 1,671,996 | North African and Middle Eastern | | North and Latin American | | Pasifika | Sub-Saharan African |
|---|---|---|---|---|---|---|---|
| | | Jewish N = 1,842 | Other North African and Middle Eastern N = 21,814 | North American N = 2,305 | Other North and Latin American N = 4,255 | Pasifika N = 2,299 | Sub-Saharan African N = 4,712 |
| **Age, median (IQR)** | 80 (77-85) | 81 (77-86) | 80 (77-84) | 78 (76-82) | 79 (77-83) | 79 (76-82) | 80 (77-84) |
| **Sex, n(%)** | | | | | | | |
| Female | 920,562 (55.1) | 996 (54.1) | 10,471 (48.0) | 1,125 (48.8) | 2,442 (57.4) | 1,353 (58.9) | 2,709 (57.5) |
| Male | 751,434 (44.9) | 846 (45.9) | 11,343 (52.0) | 1,180 (51.2) | 1,813 (42.6) | 946 (41.1) | 2,003 (42.5) |
| **Country of birth, n(%)[a]** | | | | | | | |
| Born in Australia | 998,596 (61.3) | 263 (14.6) | 856 (4.0) | 709 (31.3) | 58 (1.4) | 107 (4.8) | 207 (4.5) |
| Born Overseas | 631,735 (38.7) | 1,541 (85.4) | 20,606 (96.0) | 1,558 (68.7) | 4,110 (98.6) | 2,143 (95.2) | 4,412 (95.5) |
| **English proficiency, n(%)[a]** | | | | | | | |
| Not well/Not at all | 121,888 (7.4) | 445 (24.4) | 10,538 (48.6) | n/a[b] | 1,703 (40.4) | 633 (27.8) | 432 (9.2) |
| Very well/Well | 1,530,648 (92.6) | 1,382 (75.6) | 11,153 (51.4) | n/a[b] | 2,513 (59.6) | 1,642 (72.2) | 4,247 (90.8) |
| **Education, n(%)[a]** | | | | | | | |
| No bachelor's degree | 1,338,548 (88.1) | 949 (56.2) | 18,214 (87.8) | 1,348 (62.7) | 3,504 (90.1) | 1,941 (93.9) | 3,589 (84.2) |
| Postgraduate/Bachelor degree | 181,538 (11.9) | 740 (43.8) | 2,538 (12.2) | 802 (37.3) | 383 (9.9) | 127 (6.1) | 676 (15.8) |
| **SEIFA, n(%)[a]** | | | | | | | |
| Less disadvantage | 472,500 (28.3) | 1,312 (71.2) | 4,638 (21.3) | 825 (35.8) | 890 (20.9) | 357 (15.5) | 1,610 (34.2) |
| Moderate disadvantage | 696,098 (41.7) | 409 (22.2) | 6,814 (31.2) | 888 (38.6) | 1,588 (37.3) | 825 (35.9) | 1,787 (37.9) |
| Greater disadvantage | 502,491 (30.1) | 121 (6.6) | 10,358 (47.5) | 590 (25.6) | 1,774 (41.7) | 1,116 (48.6) | 1,312 (27.9) |
| **Remoteness, n(%)[a]** | | | | | | | |
| Major cities | 1,134,399 (67.9) | 1,760 (95.5) | 21,080 (96.7) | 1,592 (69.1) | 3,970 (93.4) | 1,974 (85.9) | 4,109 (87.3) |
| Non-major cities | 536,718 (32.1) | 82 (4.5) | 730 (3.3) | 711 (30.9) | 282 (6.6) | 324 (14.1) | 600 (12.7) |
| **Nursing home, n(%)[a]** | | | | | | | |
| Not in nursing home | 1,596,895 (95.5) | 1,813 (98.4) | 21,263 (97.5) | 2,187 (94.9) | 4,132 (97.1) | 2,202 (95.8) | 4,464 (94.7) |
| Nursing home | 75,099 (4.5) | 29 (1.6) | 551 (2.5) | 118 (5.1) | 123 (2.9) | 97 (4.2) | 248 (5.3) |
| **Long-term health condition, n(%)[a]** | | | | | | | |
| No condition | 394,795 (24.4) | 419 (23.3) | 4,702 (22.1) | 617 (27.3) | 1,115 (26.9) | 499 (22.5) | 1,272 (27.9) |
| One or more condition(s) | 1,225,941 (75.6) | 1,383 (76.7) | 16,598 (77.9) | 1,647 (72.7) | 3,024 (73.1) | 1,722 (77.5) | 3,295 (72.1) |

Abbreviations: ABS: Australian Bureau of Statistics; IQR: Interquartile range; SEIFA: Socio-Economic Indexes for Areas (Index of Relative Socio-economic Disadvantage (IRSD) based on Statistical Area Level 2 (SA2)). [a] Percentages are calculated based on valid responses (excluding missing values). Proportion of missing data: Education (9.1%), Long-term health conditions (3.1%), and Country of birth (2.5%). Missing data were <1% for all other variables. [b] n/a: Not available. In accordance with ABS confidentiality rules, cell counts fewer than 10 have been suppressed to prevent potential re-identification.

#### Section 3. Supplementary Figures

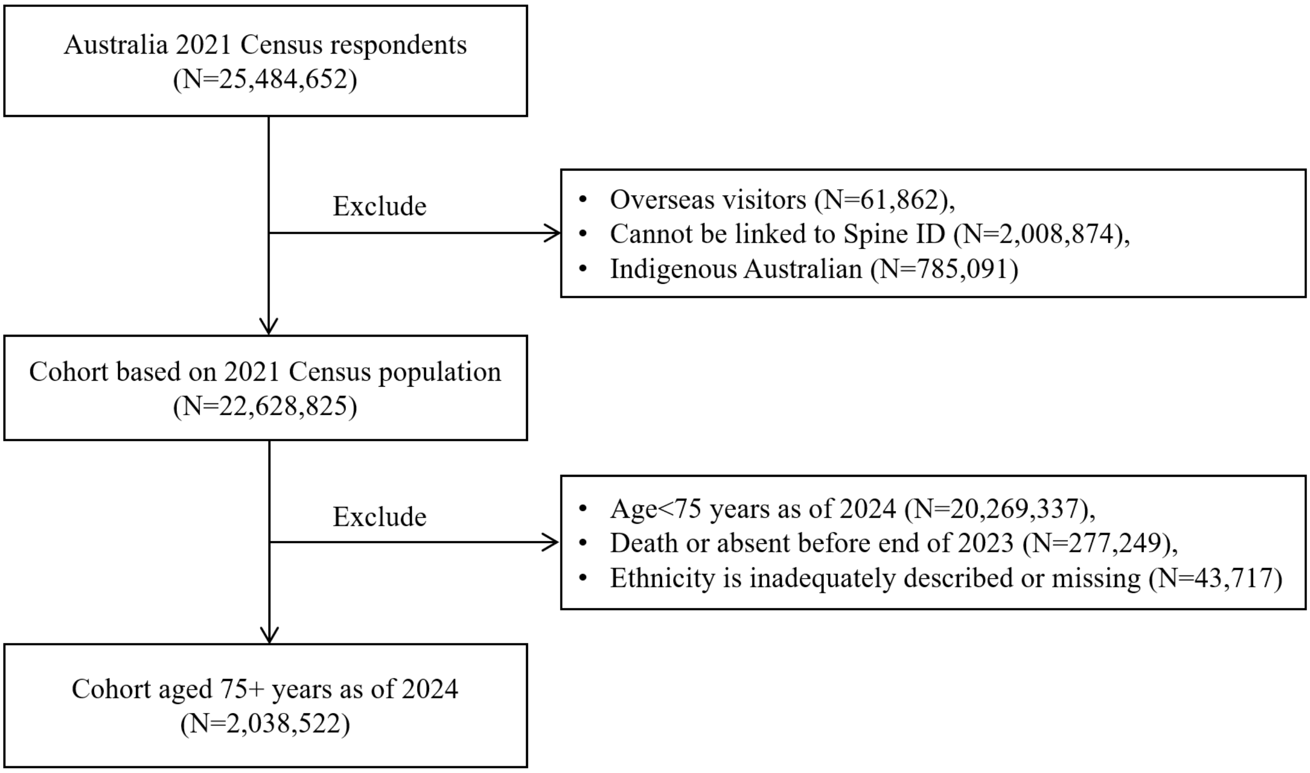

**Figure S1. Study Population Exclusion Flowchart for the Primary analysis (2024 cohort)**

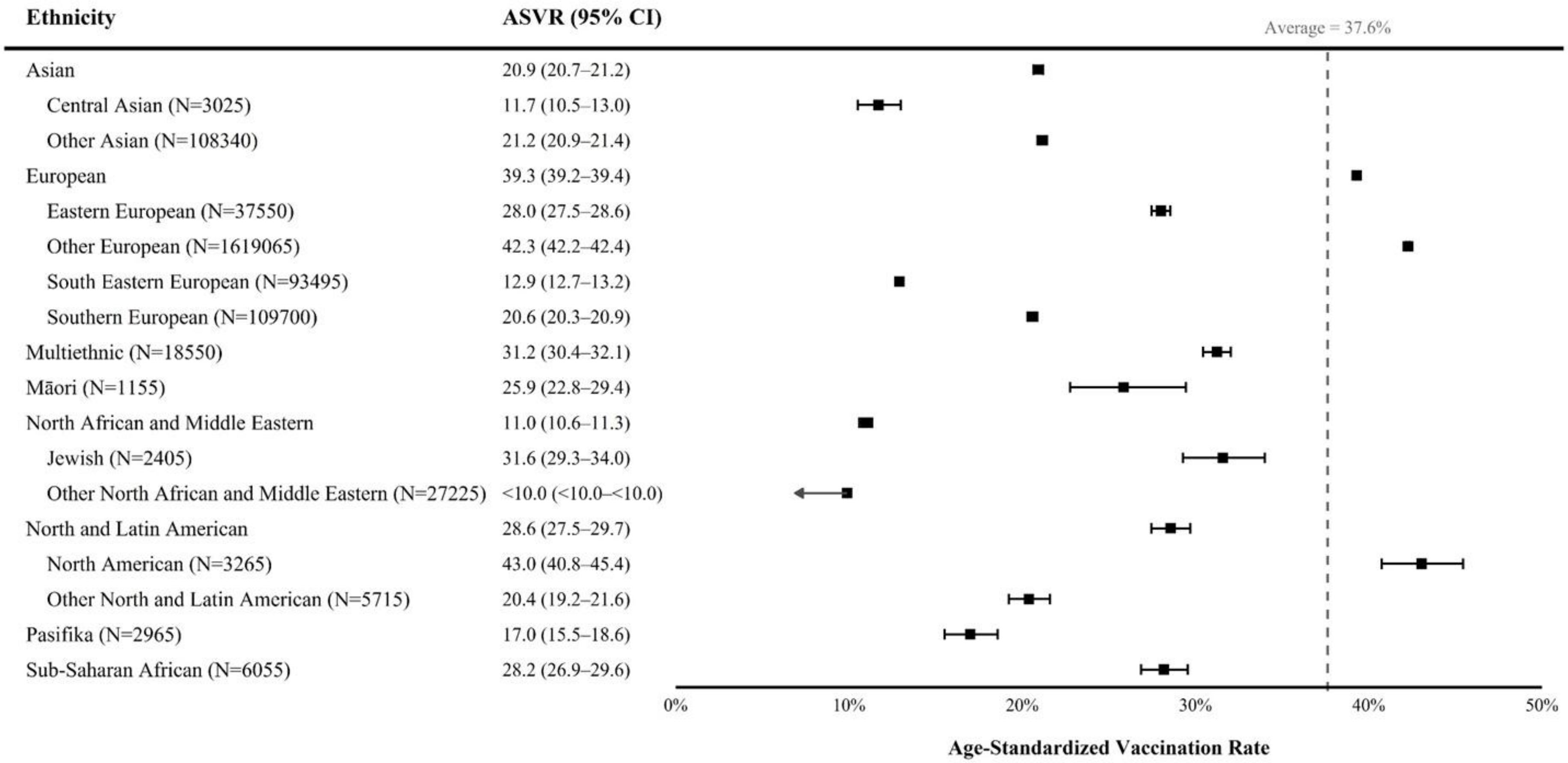

**Figure S2. Age-standardized Uptake of COVID-19 Vaccine Dose by Ethnicity, 01 January–30 September 2024.**

**Note.** In accordance with Australian Bureau of Statistics confidentiality rules, percentages below 10% were displayed as "<10.0". COVID-19 vaccination data were probabilistically rounded to the nearest multiple of five. Rates are age-standardized to the World Health Organization standard population. N: number of individuals in this ethnic group, with counts probabilistically rounded to the nearest multiple of five; ASVR: Age-standardized vaccination rate (%); 95% CI: 95% Confidence Interval.

| Ethnicity | Education | N | ASVR (95% CI) |
|---|---|---|---|
| Asian | No bachelor's degree | 77190 | 15.3 (15.0-15.6) |
| | Postgraduate/Bachelor degree | 28750 | 21.0 (20.4-21.5) |
| Central Asian | No bachelor's degree | 2430 | <10.0 (<10.0-10.3) |
| | Postgraduate/Bachelor degree | 440 | 14.0 (10.7-18.1) |
| European | No bachelor's degree | 1500215 | 31.4 (31.3-31.5) |
| | Postgraduate/Bachelor degree | 212240 | 43.1 (42.8-43.4) |
| Eastern European | No bachelor's degree | 26880 | 21.7 (21.1-22.2) |
| | Postgraduate/Bachelor degree | 7620 | 29.5 (28.2-30.7) |
| South Eastern European | No bachelor's degree | 85475 | <10.0 (<10.0-10.0) |
| | Postgraduate/Bachelor degree | 2765 | 25.3 (23.5-27.3) |
| Southern European | No bachelor's degree | 98830 | 16.4 (16.1-16.7) |
| | Postgraduate/Bachelor degree | 3205 | 30.9 (29.0-33.0) |
| Multiethnic | No bachelor's degree | 14070 | 24.8 (24.0-25.7) |
| | Postgraduate/Bachelor degree | 3380 | 32.9 (30.9-34.9) |
| Māori | No bachelor's degree | 995 | 20.0 (17.2-23.3) |
| | Postgraduate/Bachelor degree | 50 | 28.2 (15.8-48.6) |
| North African and Middle Eastern | No bachelor's degree | 23765 | <10.0 (<10.0-<10.0) |
| | Postgraduate/Bachelor degree | 4505 | 18.9 (17.7-20.3) |
| Jewish | No bachelor's degree | 1190 | 25.6 (22.6-28.8) |
| | Postgraduate/Bachelor degree | 1055 | 28.2 (25.0-31.8) |
| North and Latin American | No bachelor's degree | 6570 | 19.9 (18.8-21.0) |
| | Postgraduate/Bachelor degree | 1750 | 36.6 (33.7-39.7) |
| North American | No bachelor's degree | 1900 | 30.5 (28.0-33.2) |
| | Postgraduate/Bachelor degree | 1180 | 42.8 (39.0-46.9) |
| Pasifika | No bachelor's degree | 2505 | 12.5 (11.1-14.0) |
| | Postgraduate/Bachelor degree | 170 | 20.8 (14.4-29.2) |
| Sub-Saharan African | No bachelor's degree | 4625 | 21.2 (19.9-22.6) |
| | Postgraduate/Bachelor degree | 940 | 31.5 (28.0-35.3) |

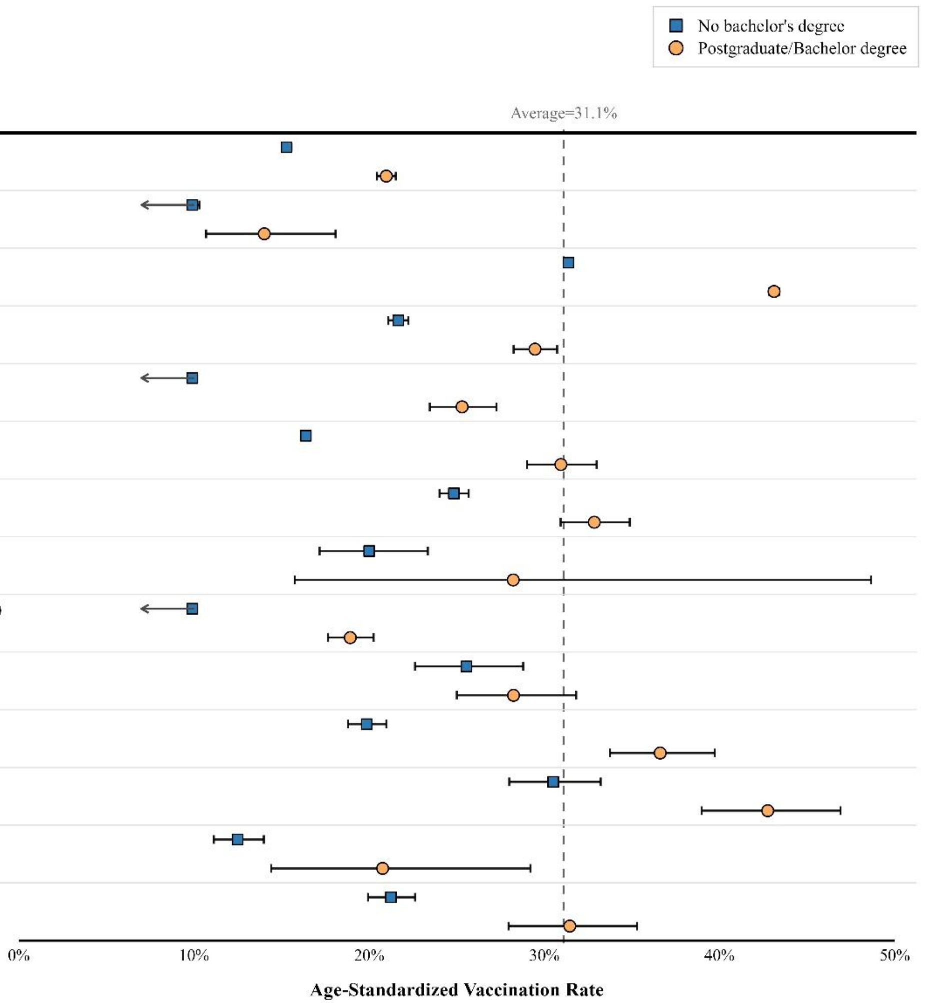

**Figure S3. Age-standardized Uptake of COVID-19 Vaccine Dose by Ethnicity and Education, 01 January-30 June 2024**

**Note.** In accordance with ABS confidentiality rules, percentages below 10% were displayed as “<10.0”. COVID-19 vaccination data were probabilistically rounded to the nearest multiple of five. Rates are age-standardized to the World Health Organization standard population. N: number of individuals in this ethnic group, with counts probabilistically rounded to the nearest multiple of five; ASVR: Age-standardized vaccination rate (%); 95% CI: 95% Confidence Interval.

| Ethnicity | Health Condition | N | ASVR (95% CI) |
|---|---|---|---|
| Asian | No condition | 37495 | 14.9 (14.5-15.3) |
| | One or more condition(s) | 71465 | 18.0 (17.7-18.4) |
| Central Asian | No condition | 920 | <10.0 (<10.0-<10.0) |
| | One or more condition(s) | 2045 | 11.0 (<10.0-12.6) |
| European | No condition | 507365 | 30.2 (30.0-30.3) |
| | One or more condition(s) | 1294355 | 33.7 (33.6-33.8) |
| Eastern European | No condition | 11655 | 21.1 (20.2-21.9) |
| | One or more condition(s) | 24845 | 24.3 (23.7-25.0) |
| South Eastern European | No condition | 28060 | <10.0 (<10.0-<10.0) |
| | One or more condition(s) | 62765 | 11.2 (10.9-11.5) |
| Southern European | No condition | 30815 | 14.9 (14.5-15.4) |
| | One or more condition(s) | 74835 | 17.8 (17.5-18.2) |
| Multiethnic | No condition | 4730 | 23.4 (22.0-24.8) |
| | One or more condition(s) | 13405 | 27.2 (26.3-28.1) |
| Māori | No condition | 275 | 14.7 (10.6-20.8) |
| | One or more condition(s) | 835 | 23.4 (20.1-27.3) |
| North African and Middle Eastern | No condition | 7405 | <10.0 (<10.0-<10.0) |
| | One or more condition(s) | 21510 | <10.0 (<10.0-<10.0) |
| Jewish | No condition | 660 | 25.6 (21.8-29.9) |
| | One or more condition(s) | 1700 | 27.5 (24.9-30.3) |
| North and Latin American | No condition | 2720 | 22.8 (20.9-24.8) |
| | One or more condition(s) | 6035 | 23.5 (22.3-24.8) |
| North American | No condition | 1000 | 33.5 (29.8-37.5) |
| | One or more condition(s) | 2200 | 36.2 (33.6-38.8) |
| Pasifika | No condition | 780 | 11.1 (<10.0-13.9) |
| | One or more condition(s) | 2085 | 13.2 (11.6-14.8) |
| Sub-Saharan African | No condition | 1915 | 21.5 (19.4-23.7) |
| | One or more condition(s) | 3945 | 23.6 (22.1-25.2) |

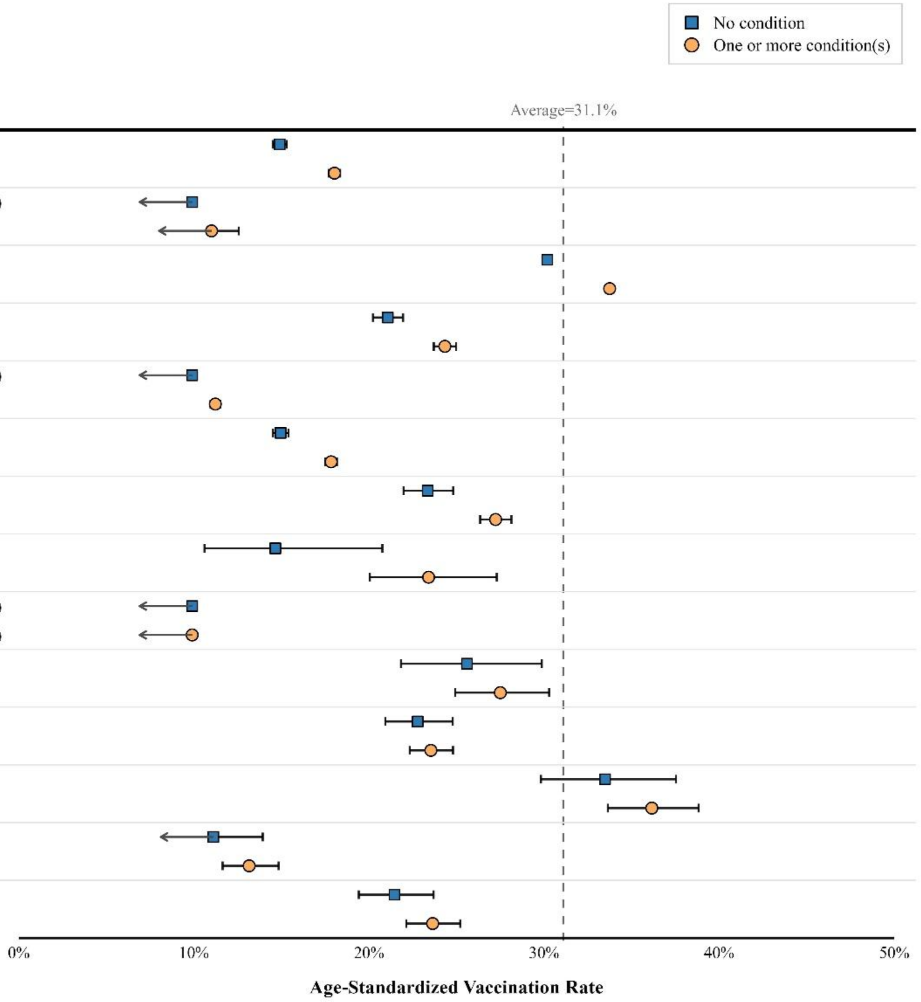

**Figure S4. Age-standardized Uptake of COVID-19 Vaccine Dose by Ethnicity and Self-reported Long-term Health Condition Status, 01 January-30 June 2024**

**Note.** In accordance with ABS confidentiality rules, percentages below 10% were displayed as “<10.0”. COVID-19 vaccination data were probabilistically rounded to the nearest multiple of five. Rates are age-standardized to the World Health Organization standard population. N: number of individuals in this ethnic group, with counts probabilistically rounded to the nearest multiple of five; ASVR: Age-standardized vaccination rate (%); 95% CI: 95% Confidence Interval.

| Ethnicity | SEIFA | N | ASVR (95% CI) |
|---|---|---|---|
| Asian | Less disadvantage | 39030 | 20.5 (20.1-21.0) |
| | Moderate disadvantage | 40885 | 16.4 (16.0-16.8) |
| | Greater disadvantage | 31410 | 13.0 (12.6-13.5) |
| Central Asian | Less disadvantage | 1225 | 14.6 (12.5-16.9) |
| | Moderate disadvantage | 1025 | <10.0 (<10.0-<10.0) |
| | Greater disadvantage | 790 | <10.0 (<10.0-<10.0) |
| European | Less disadvantage | 522480 | 37.7 (37.5-37.9) |
| | Moderate disadvantage | 786530 | 32.2 (32.1-32.3) |
| | Greater disadvantage | 549315 | 28.3 (28.1-28.4) |
| Eastern European | Less disadvantage | 14680 | 27.7 (26.9-28.6) |
| | Moderate disadvantage | 13890 | 22.1 (21.4-23.0) |
| | Greater disadvantage | 8960 | 16.8 (15.9-17.7) |
| South Eastern European | Less disadvantage | 27520 | 13.7 (13.3-14.2) |
| | Moderate disadvantage | 40100 | 10.1 (<10.0-10.4) |
| | Greater disadvantage | 25850 | <10.0 (<10.0-<10.0) |
| Southern European | Less disadvantage | 31015 | 20.4 (19.9-21.0) |
| | Moderate disadvantage | 46835 | 16.5 (16.1-16.9) |
| | Greater disadvantage | 31825 | 13.9 (13.4-14.3) |
| Multiethnic | Less disadvantage | 6030 | 31.8 (30.4-33.3) |
| | Moderate disadvantage | 7220 | 25.4 (24.2-26.6) |
| | Greater disadvantage | 5275 | 20.5 (19.3-21.8) |
| Māori | Less disadvantage | 170 | 24.5 (17.9-33.5) |
| | Moderate disadvantage | 505 | 18.7 (15.1-23.4) |
| | Greater disadvantage | 475 | 17.6 (14.0-23.1) |
| North African and Middle Eastern | Less disadvantage | 7475 | 17.5 (16.6-18.5) |
| | Moderate disadvantage | 9005 | <10.0 (<10.0-10.2) |
| | Greater disadvantage | 13145 | <10.0 (<10.0-<10.0) |
| Jewish | Less disadvantage | 1755 | 27.9 (25.4-30.7) |
| | Moderate disadvantage | 520 | 23.0 (18.8-27.8) |
| | Greater disadvantage | 155 | 24.8 (17.2-34.7) |
| North and Latin American | Less disadvantage | 2405 | 32.9 (30.6-35.3) |
| | Moderate disadvantage | 3450 | 22.1 (20.6-23.8) |
| | Greater disadvantage | 3105 | 16.9 (15.5-18.4) |
| North American | Less disadvantage | 1155 | 43.5 (39.7-47.6) |
| | Moderate disadvantage | 1295 | 30.9 (27.8-34.2) |
| | Greater disadvantage | 810 | 30.8 (27.0-35.0) |
| Pasifika | Less disadvantage | 445 | 23.4 (19.1-28.5) |
| | Moderate disadvantage | 1075 | 13.8 (11.7-16.3) |
| | Greater disadvantage | 1440 | 10.0 (<10.0-11.9) |
| Sub-Saharan African | Less disadvantage | 2075 | 30.2 (27.8-32.6) |
| | Moderate disadvantage | 2345 | 20.9 (19.1-22.8) |
| | Greater disadvantage | 1635 | 16.7 (14.8-18.8) |

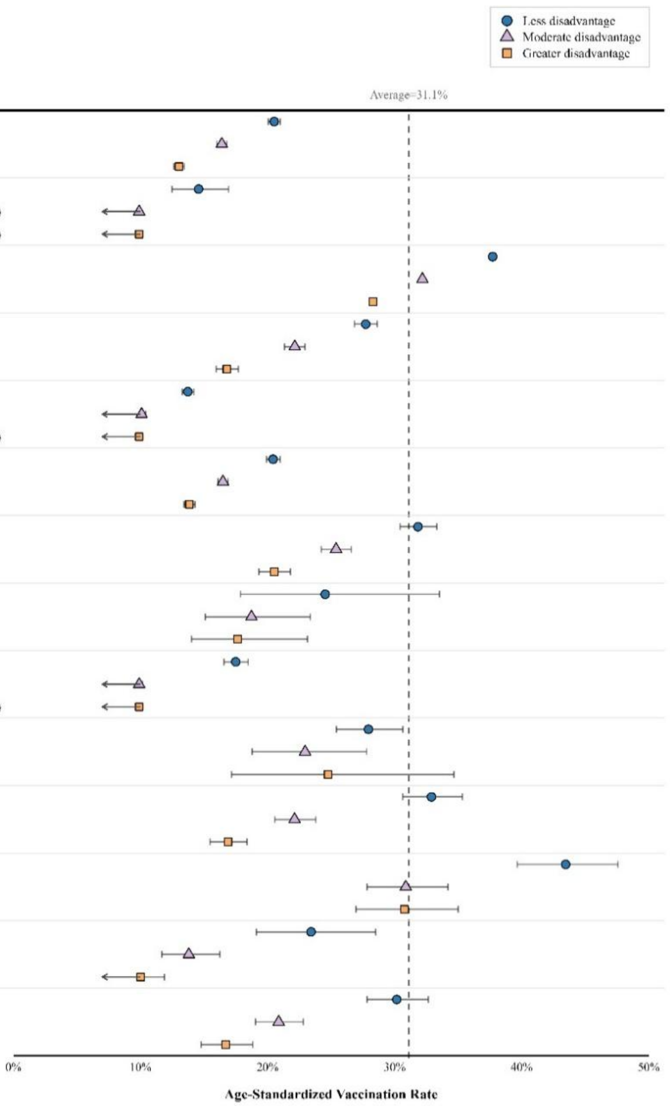

**Figure S5. Age-standardized Uptake of COVID-19 Vaccine Dose by Ethnicity and SEIFA, 01 January-30 June 2024**

**Note.** In accordance with ABS confidentiality rules, percentages below 10% were displayed as “<10.0”. COVID-19 vaccination data were probabilistically rounded to the nearest multiple of five. Rates are age-standardized to the World Health Organization standard population. SEIFA: Socio-Economic Indexes for Areas (Index of Relative Socio-economic Disadvantage (IRSD) based on Statistical Area Level 2 (SA2)); N: number of individuals in this ethnic group, with counts probabilistically rounded to the nearest multiple of five; ASVR: Age-standardized vaccination rate (%); 95% CI: 95% Confidence Interval.

| Ethnicity | Remoteness | N | ASVR (95% CI) |
|---|---|---|---|
| Asian | Major cities | 106120 | 16.7 (16.4-16.9) |
| | Non-major cities | 5225 | 21.1 (19.8-22.4) |
| Central Asian | Major cities | 2945 | <10.0 (<10.0-10.8) |
| | Non-major cities | 75 | 27.4 (16.3-44.2) |
| European | Major cities | 1203520 | 32.6 (32.5-32.7) |
| | Non-major cities | 654840 | 32.6 (32.4-32.7) |
| Eastern European | Major cities | 31475 | 22.7 (22.1-23.2) |
| | Non-major cities | 6050 | 24.8 (23.5-26.1) |
| South Eastern European | Major cities | 86970 | 10.0 (<10.0-10.2) |
| | Non-major cities | 6500 | 16.8 (15.8-17.9) |
| Southern European | Major cities | 93910 | 16.3 (16.0-16.6) |
| | Non-major cities | 15750 | 19.8 (19.1-20.6) |
| Multiethnic | Major cities | 15155 | 25.4 (24.6-26.2) |
| | Non-major cities | 3370 | 29.3 (27.5-31.3) |
| Māori | Major cities | 845 | 20.7 (17.6-24.3) |
| | Non-major cities | 325 | 23.2 (18.1-29.5) |
| North African and Middle Eastern | Major cities | 28610 | <10.0 (<10.0-<10.0) |
| | Non-major cities | 1030 | 20.8 (18.1-23.8) |
| Jewish | Major cities | 2280 | 26.6 (24.4-28.9) |
| | Non-major cities | 120 | 37.7 (27.4-50.8) |
| North and Latin American | Major cities | 7485 | 21.8 (20.7-22.9) |
| | Non-major cities | 1490 | 29.2 (26.4-32.2) |
| North American | Major cities | 2180 | 36.3 (33.8-39.0) |
| | Non-major cities | 1075 | 32.2 (28.8-36.0) |
| Pasifika | Major cities | 2520 | 12.8 (11.5-14.4) |
| | Non-major cities | 435 | 19.2 (15.1-24.5) |
| Sub-Saharan African | Major cities | 5250 | 22.7 (21.4-24.0) |
| | Non-major cities | 815 | 24.1 (20.8-27.7) |

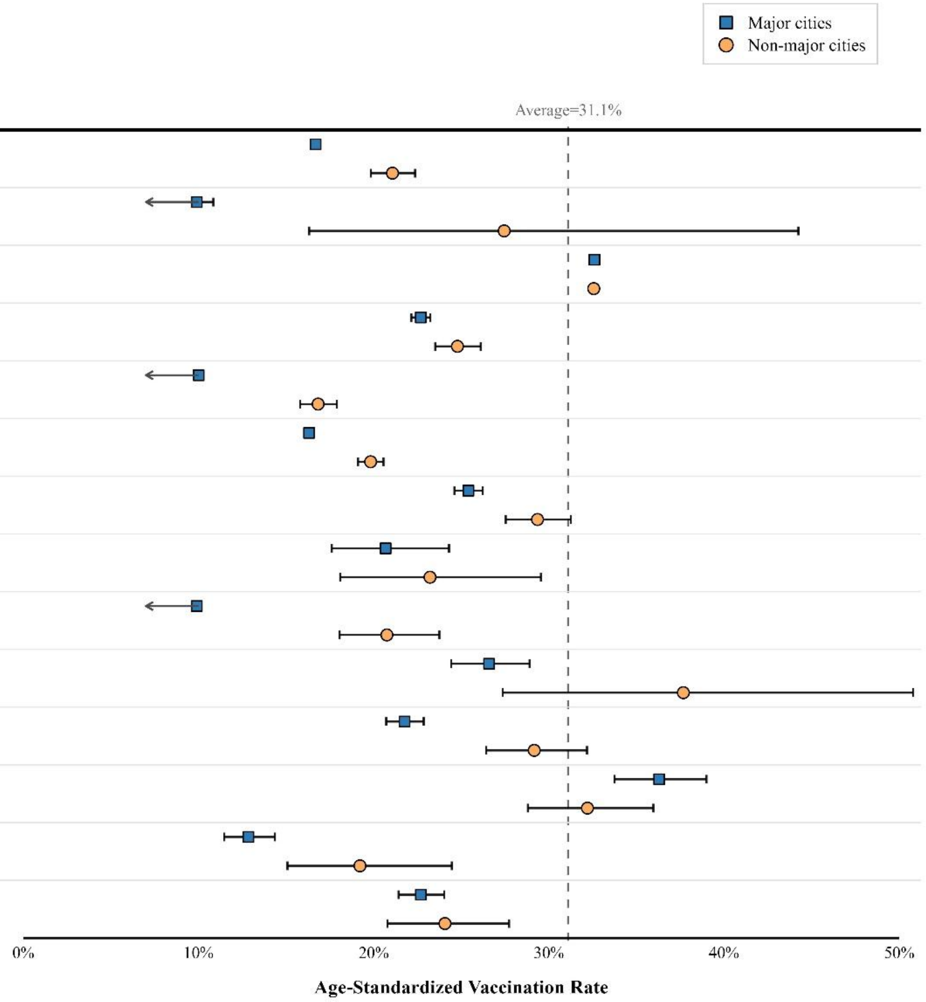

**Figure S6. Age-standardized Uptake of COVID-19 Vaccine Dose by Ethnicity and Remoteness, 01 January-30 June 2024**

**Note.** In accordance with ABS confidentiality rules, percentages below 10% were displayed as "<10.0". COVID-19 vaccination data were probabilistically rounded to the nearest multiple of five. Rates are age-standardized to the World Health Organization standard population. N: number of individuals in this ethnic group, with counts probabilistically rounded to the nearest multiple of five; ASVR: Age-standardized vaccination rate (%); 95% CI: 95% Confidence Interval.

| Ethnicity | Nursing Home | N | ASVR (95% CI) |
|---|---|---|---|
| Asian | Not in nursing home | 109980 | 16.7 (16.5-17.0) |
| | Nursing home | 1385 | 35.3 (30.2-41.1) |
| European | Not in nursing home | 1824365 | 32.5 (32.4-32.6) |
| | Nursing home | 35460 | 38.6 (37.5-39.6) |
| Eastern European | Not in nursing home | 36840 | 22.9 (22.4-23.4) |
| | Nursing home | 720 | 32.5 (25.7-40.8) |
| South Eastern European | Not in nursing home | 92040 | 10.3 (10.1-10.5) |
| | Nursing home | 1455 | 27.4 (22.9-32.7) |
| Southern European | Not in nursing home | 107635 | 16.7 (16.4-16.9) |
| | Nursing home | 2080 | 31.9 (27.2-37.2) |
| Multiethnic | Not in nursing home | 18325 | 26.0 (25.3-26.8) |
| | Nursing home | 220 | 36.0 (23.9-53.2) |
| North African and Middle Eastern | Not in nursing home | 29310 | <10.0 (<10.0-<10.0) |
| | Nursing home | 330 | 21.9 (15.7-30.4) |
| North and Latin American | Not in nursing home | 8830 | 22.9 (21.9-24.0) |
| | Nursing home | 145 | 38.4 (25.9-55.4) |
| North American | Not in nursing home | 3195 | 35.0 (32.9-37.2) |
| | Nursing home | 70 | 50.1 (30.3-79.0) |
| Pasifika | Not in nursing home | 2900 | 12.5 (11.2-13.9) |
| | Nursing home | 60 | 26.3 (13.0-48.8) |
| Sub-Saharan African | Not in nursing home | 5915 | 22.8 (21.6-24.0) |
| | Nursing home | 135 | 37.8 (24.7-56.3) |

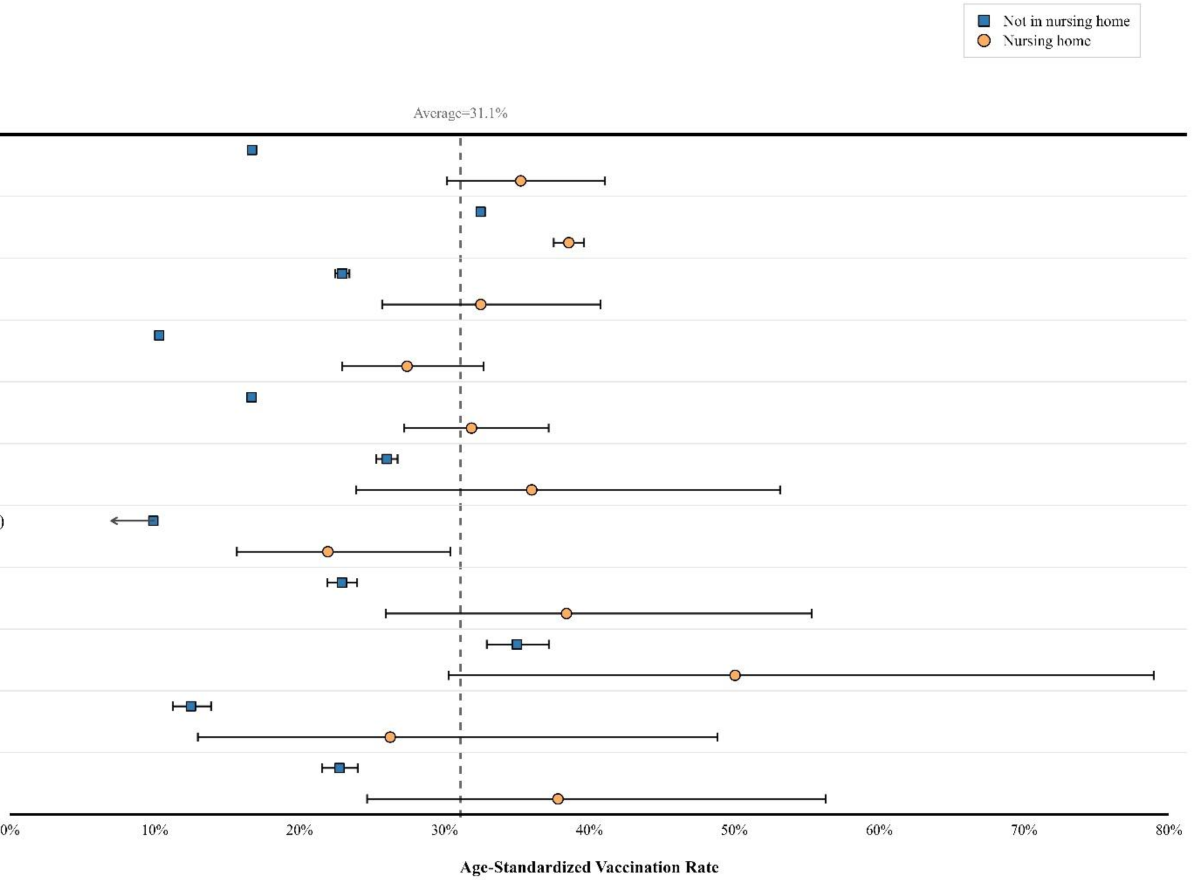

**Figure S7. Age-standardized Uptake of COVID-19 Vaccine Dose by Ethnicity and Nursing Home Residence, 01 January-30 June 2024**

**Note.** In accordance with ABS confidentiality rules, cell counts fewer than 10 were suppressed to prevent potential re-identification; therefore, the Central Asian, Māori and Jewish group are not shown in this figure. Percentages below 10% were displayed as "<10.0". COVID-19 vaccination data were probabilistically rounded to the nearest multiple of five. Rates are age-standardized to the World Health Organization standard population. N: number of individuals in this ethnic group, with counts probabilistically rounded to the nearest multiple of five; ASVR: Age-standardized vaccination rate (%); 95% CI: 95% Confidence Interval.

| Ethnicity | Sex | N | ASVR (95% CI) |
|---|---|---|---|
| Asian | Male | 49700 | 17.1 (16.7-17.4) |
| | Female | 61670 | 16.8 (16.5-17.1) |
| Central Asian | Male | 1440 | 10.1 (<10.0-11.9) |
| | Female | 1590 | <10.0 (<10.0-10.3) |
| European | Male | 841855 | 32.8 (32.7-32.9) |
| | Female | 1017955 | 32.4 (32.2-32.5) |
| Eastern European | Male | 16475 | 24.2 (23.4-24.9) |
| | Female | 21075 | 22.1 (21.5-22.8) |
| South Eastern European | Male | 42970 | 11.2 (10.9-11.5) |
| | Female | 50525 | <10.0 (<10.0-10.2) |
| Southern European | Male | 51495 | 17.9 (17.6-18.3) |
| | Female | 58200 | 15.7 (15.4-16.1) |
| Multiethnic | Male | 8195 | 26.7 (25.6-27.9) |
| | Female | 10350 | 25.5 (24.6-26.5) |
| Māori | Male | 570 | 21.2 (17.4-25.9) |
| | Female | 575 | 22.8 (18.7-27.9) |
| North African and Middle Eastern | Male | 15190 | <10.0 (<10.0-10.1) |
| | Female | 14455 | <10.0 (<10.0-<10.0) |
| Jewish | Male | 1155 | 27.3 (24.3-30.7) |
| | Female | 1255 | 26.8 (23.9-30.0) |
| North and Latin American | Male | 4100 | 23.9 (22.5-25.5) |
| | Female | 4875 | 22.4 (21.1-23.8) |
| North American | Male | 1665 | 34.9 (32.0-38.0) |
| | Female | 1580 | 36.3 (33.3-39.5) |
| Pasifika | Male | 1225 | 12.7 (10.8-14.9) |
| | Female | 1725 | 13.9 (12.2-15.8) |
| Sub-Saharan African | Male | 2640 | 23.5 (21.7-25.5) |
| | Female | 3420 | 22.3 (20.8-24.0) |

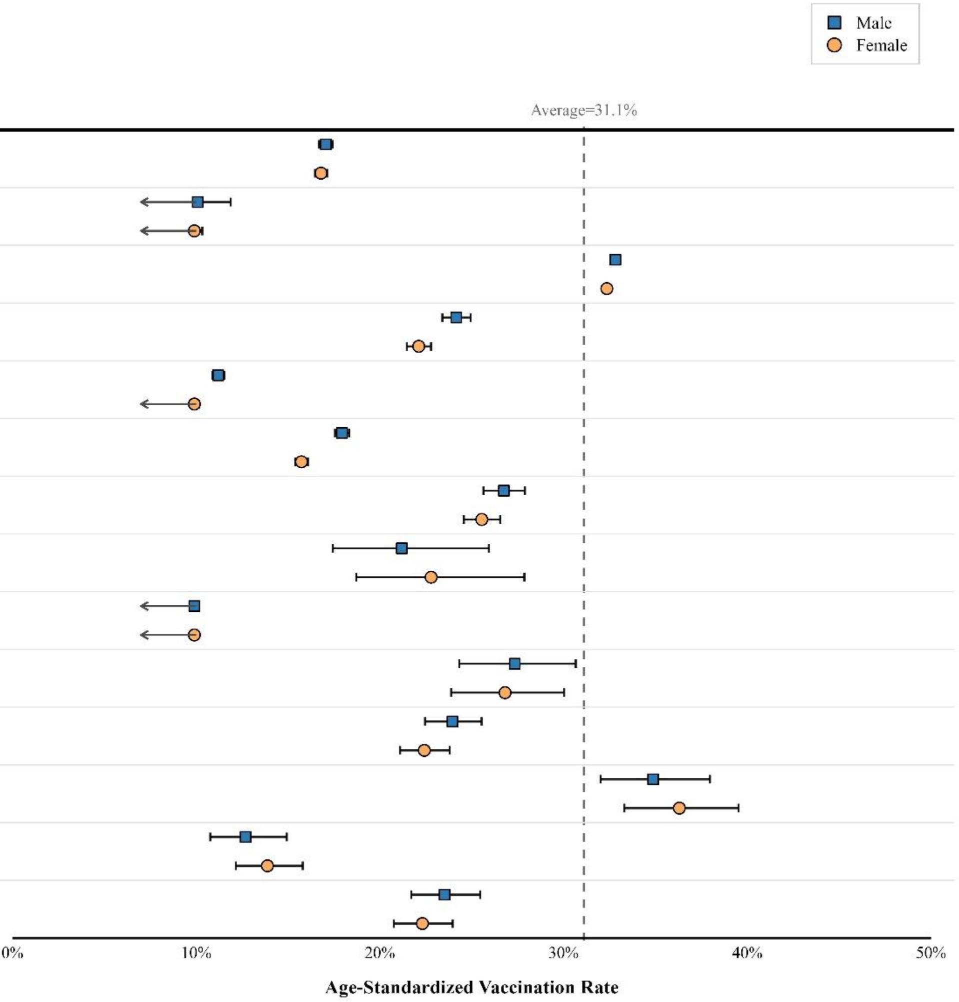

**Figure S8. Age-standardized Uptake of COVID-19 Vaccine Dose by Ethnicity and Sex, 01 January-30 June 2024**

**Note.** In accordance with ABS confidentiality rules, percentages below 10% were displayed as "<10.0". COVID-19 vaccination data were probabilistically rounded to the nearest multiple of five. Rates are age-standardized to the World Health Organization standard population. N: number of individuals in this ethnic group, with counts probabilistically rounded to the nearest multiple of five; ASVR: Age-standardized vaccination rate (%); 95% CI: 95% Confidence Interval.

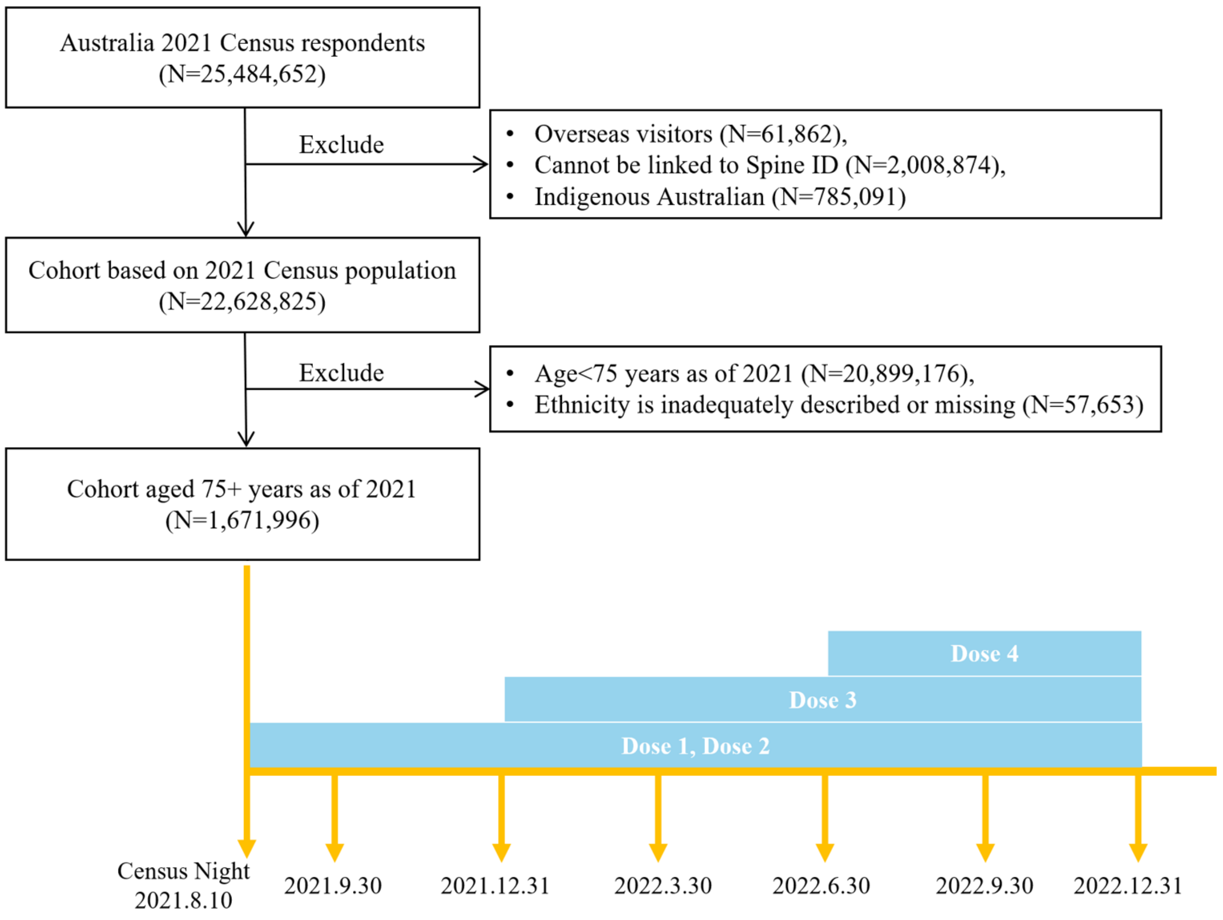

**Figure S9. Study Population Exclusion Flowchart and Study Period for Dose 1-4 in the Secondary Analysis (2021 cohort)**

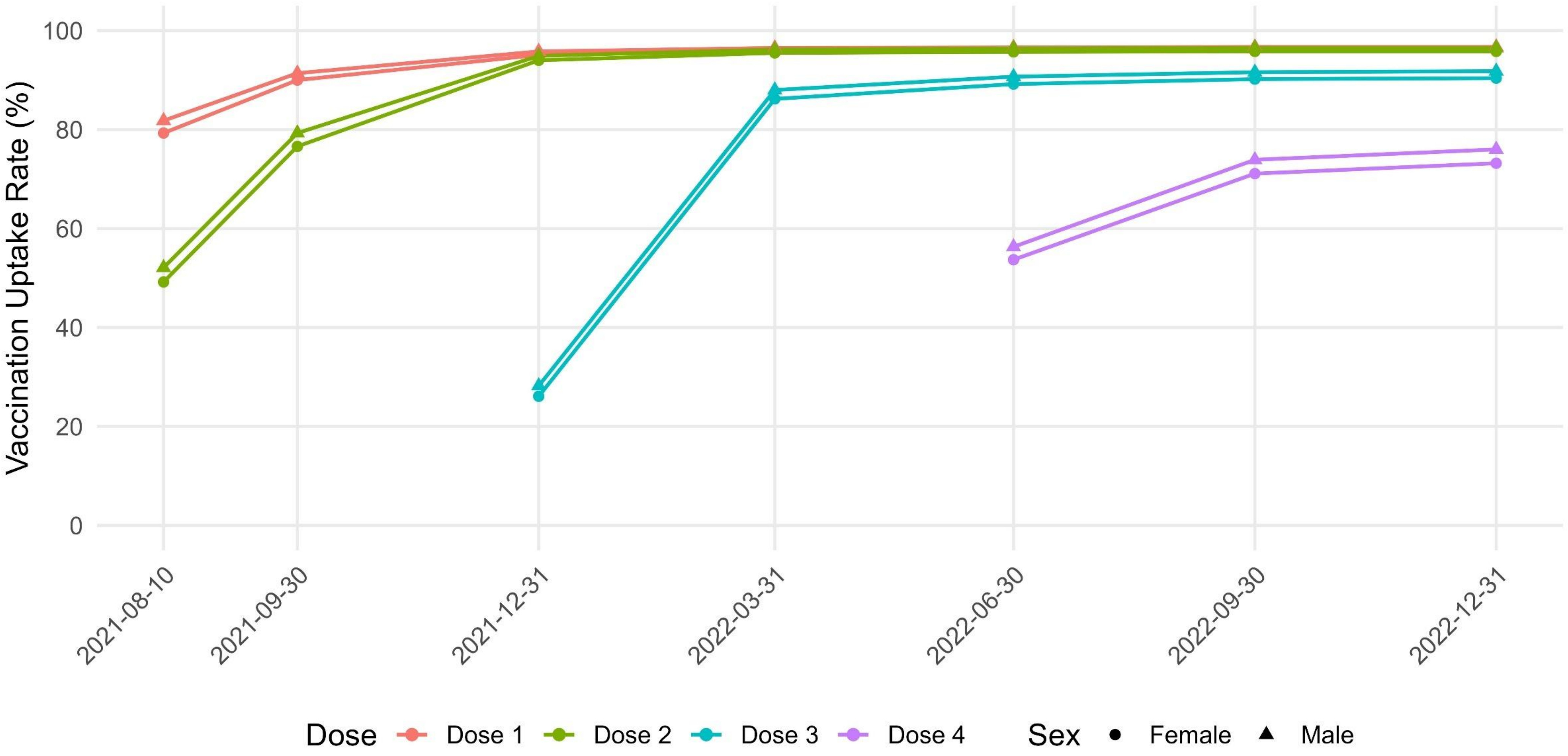

**Figure S10. Age-standardized COVID-19 Vaccination Uptake Rate by Dose and Sex**

**Note.** Rates are age-standardized to the World Health Organization standard population.

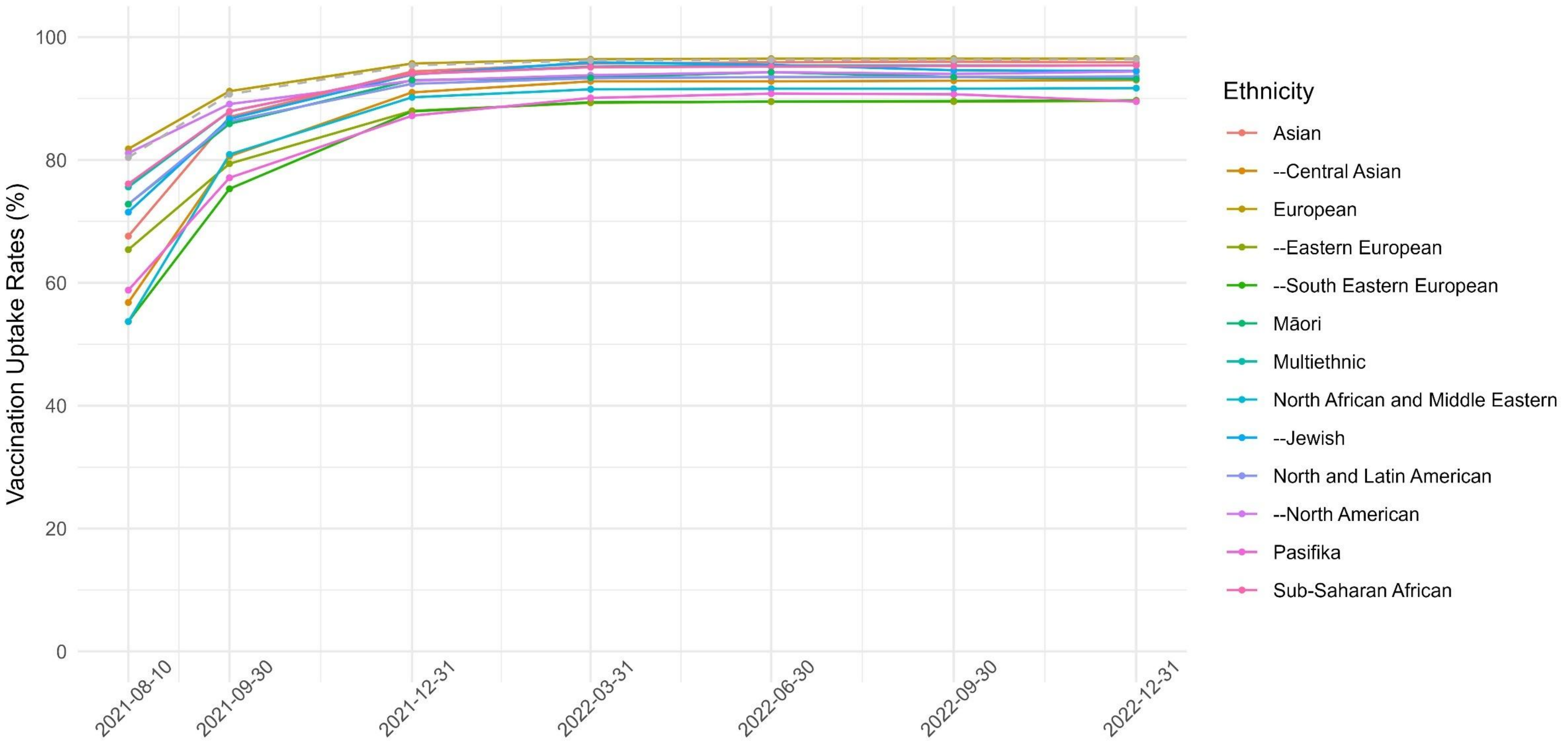

**Figure S11. Age-standardized Uptake of COVID-19 First Dose by Ethnicity, 10 August 2021-31 December 2022**

**Note.** Rates are age-standardized to the World Health Organization standard population. An interactive version of this figure, allowing users to select and compare specific ethnic groups, is available in the OSF repository: https://osf.io/wgtpk/files.

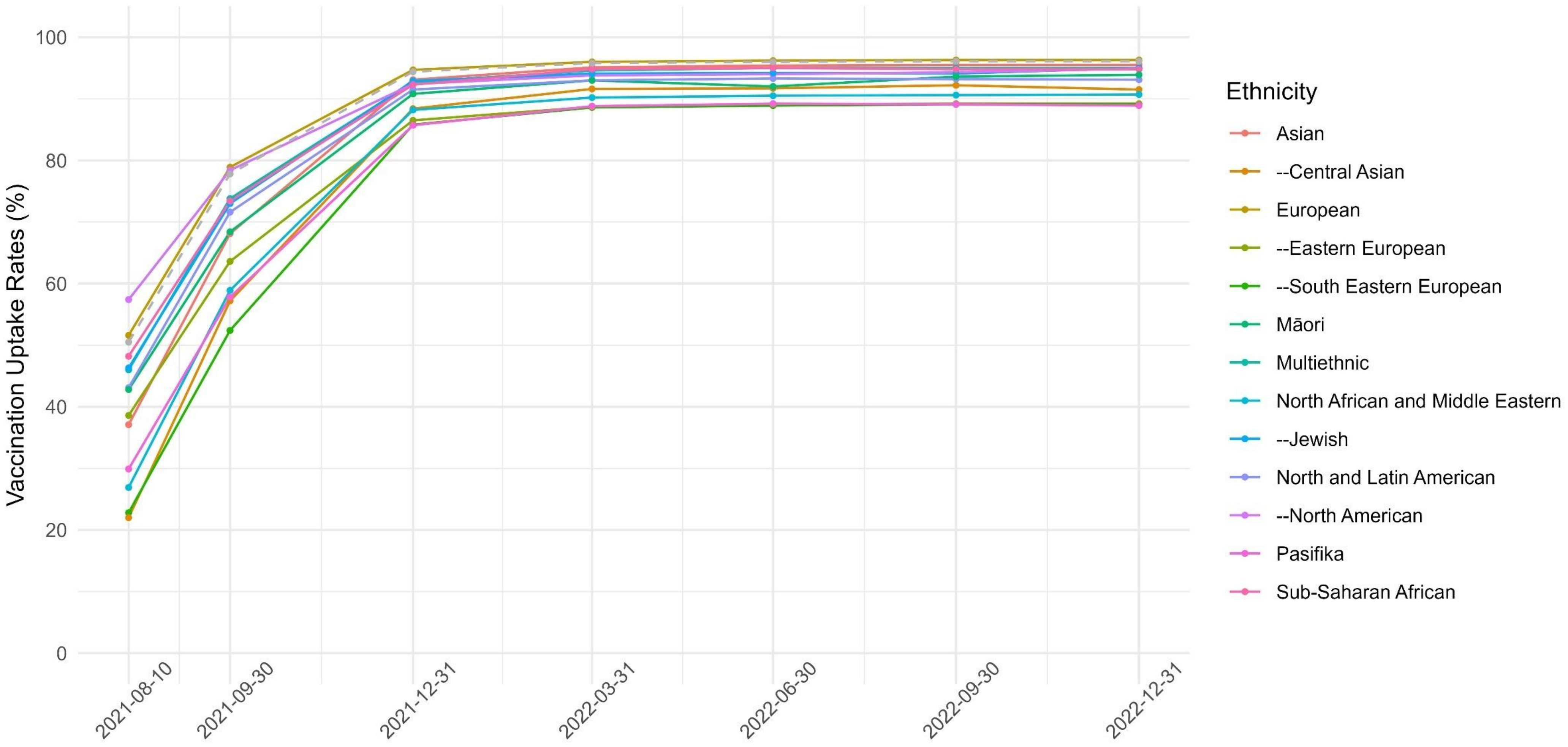

**Figure S12. Age-standardized Uptake of COVID-19 Second Dose by Ethnicity, 10 August 2021-31 December 2022**

**Note.** Rates are age-standardized to the World Health Organization standard population. An interactive version of this figure, allowing users to select and compare specific ethnic groups, is available in the OSF repository: https://osf.io/wgtpk/files.

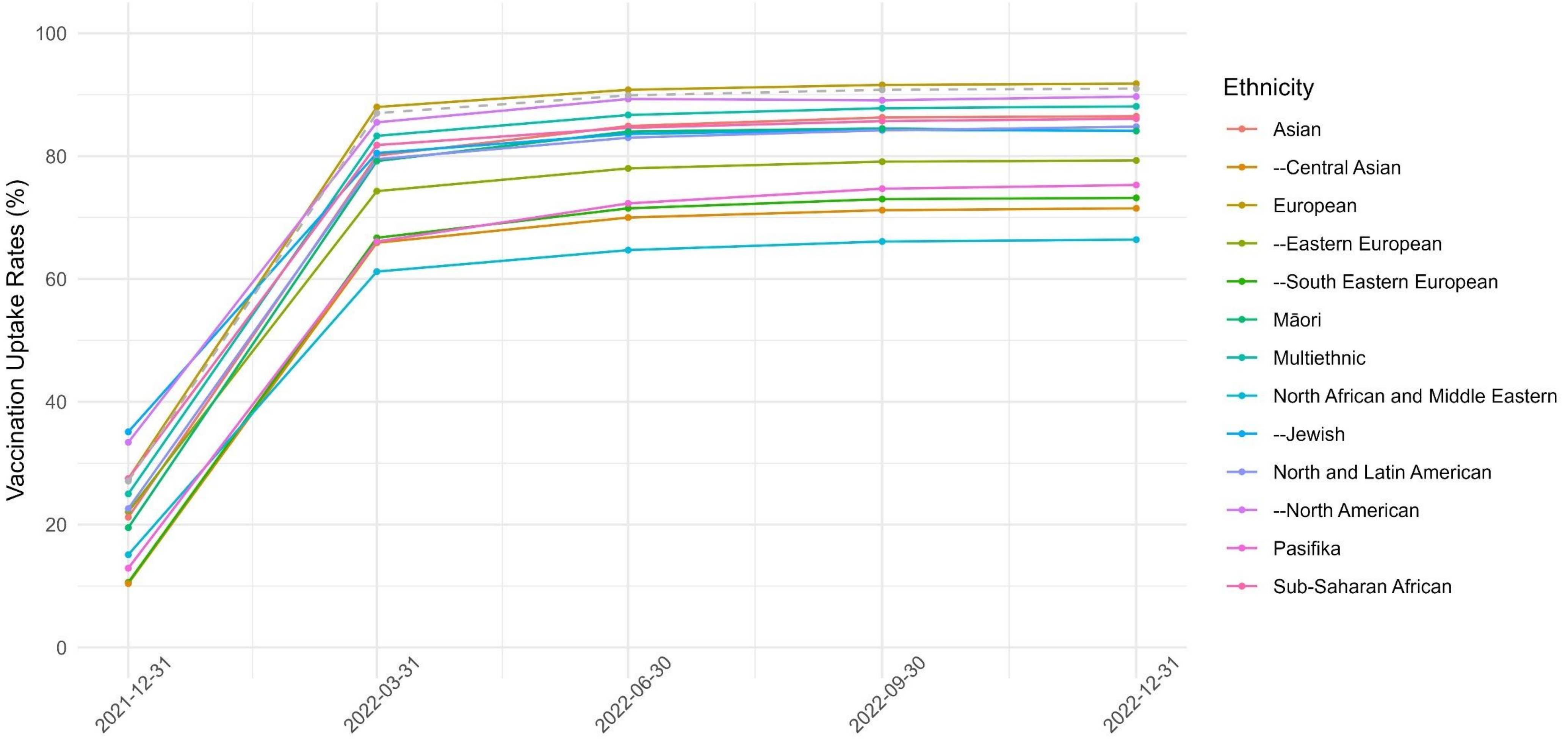

**Figure S13. Age-standardized Uptake of COVID-19 Third Dose by Ethnicity, 31 December 2021-31 December 2022**

**Note.** Rates are age-standardized to the World Health Organization standard population. An interactive version of this figure, allowing users to select and compare specific ethnic groups, is available in the OSF repository: https://osf.io/wgtpk/files.

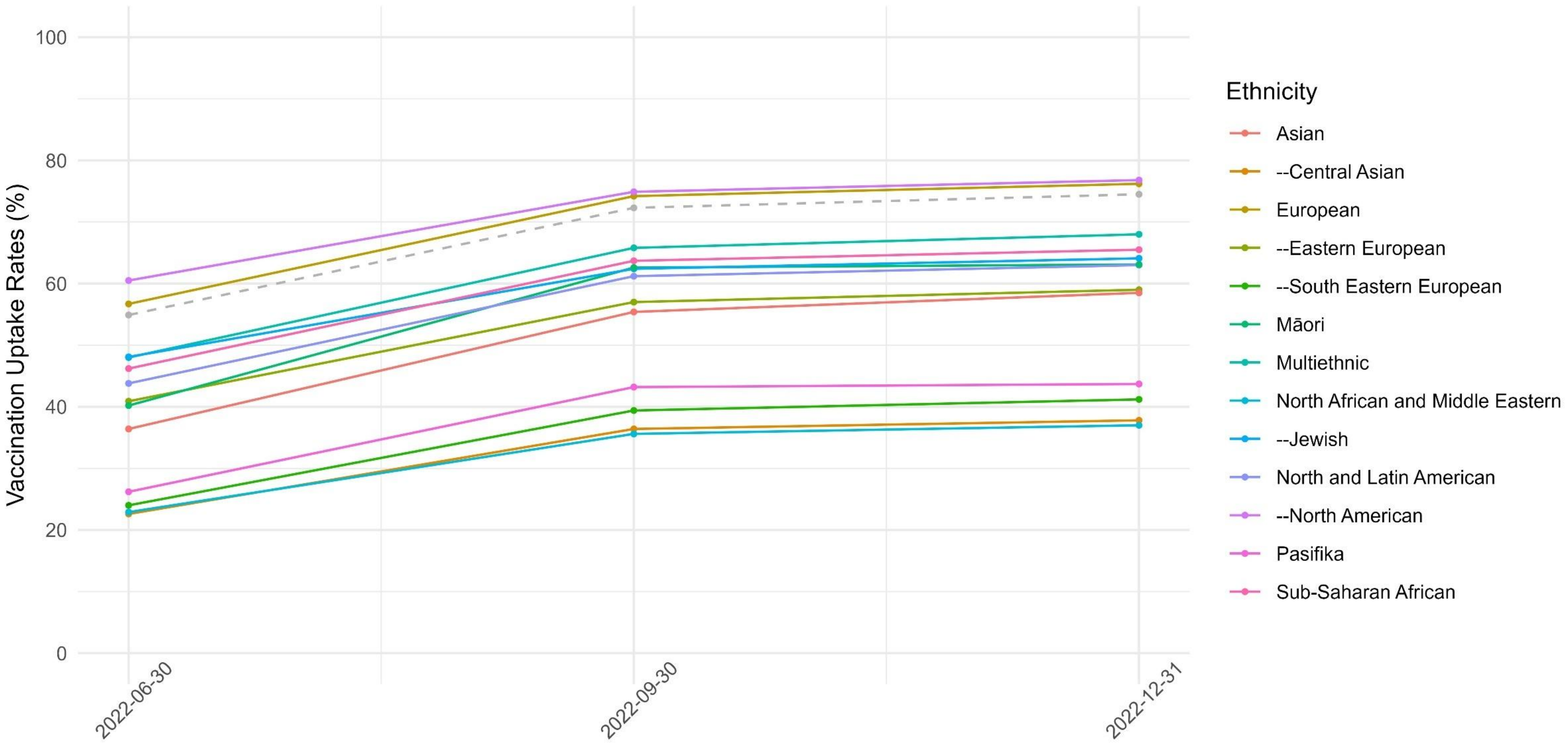

**Figure S14. Age-standardized Uptake of COVID-19 Fourth Dose by Ethnicity, 30 June 2022-31 December 2022**

**Note.** Rates are age-standardized to the World Health Organization standard population. An interactive version of this figure, allowing users to select and compare specific ethnic groups, is available in the OSF repository: https://osf.io/wgtpk/files.
